# Polygenic profiles define aspects of clinical heterogeneity in ADHD

**DOI:** 10.1101/2021.07.13.21260299

**Authors:** Sonja LaBianca, Isabell Brikell, Dorte Helenius, Robert Loughnan, Joel Mefford, Clare E. Palmer, Rebecca Walker, Jesper R. Gådin, Morten Krebs, Vivek Appadurai, Morteza Vaez, Esben Agerbo, Marianne Gørtz Pedersen, Anders D. Børglum, David M. Hougaard, Ole Mors, Merete Nordentoft, Preben Bo Mortensen, Kenneth S. Kendler, Terry L. Jernigan, Daniel H. Geschwind, Andrés Ingason, Andrew W. Dahl, Noah Zaitlen, Søren Dalsgaard, Thomas M. Werge, Andrew J. Schork

**Affiliations:** Institute of Biological Psychiatry, Mental Health Center Sct. Hans, Mental Health Services Copenhagen, Roskilde, Denmark; The Lundbeck Foundation Initiative for Integrative Psychiatric Research (iPSYCH), Copenhagen, Denmark; National Centre for Register-based Research (NCRR), Department of Economics and Business Economics, Aarhus University, Aarhus, Denmark; Department of Cognitive Science, University of California San Diego, La Jolla, CA, USA; Department of Neurology, University of California, Los Angeles, Los Angeles, CA, USA; Center for Human Development, University of California San Diego, La Jolla, CA, USA; Department of Human Genetics, University of California, Los Angeles, Los Angeles, CA, USA; Center for Autism Research and Treatment, Semel Institute, David Geffen School of Medicine, University of California, Los Angeles, Los Angeles, CA, USA; Centre for Integrated Register-based Research (CIRRAU), Aarhus University, Aarhus, Denmark; Department of Biomedicine - Human Genetics, Aarhus University, Aarhus, Denmark; Centre for Integrative Sequencing (iSEQ), Aarhus University, Aarhus, Denmark; Center for Neonatal Screening, Department for Congenital Disorders, Statens Serum Institut, Copenhagen, Denmark; Psychosis Research Unit, Aarhus University Hospital – Psychiatry, Denmark; Copenhagen Mental Health Center, Mental Health Services Capital Region of Denmark Copenhagen, Copenhagen, Denmark; Department of Clinical Medicine, Faculty of Health and Medical Sciences, University of Copenhagen, Copenhagen, Denmark; Department of Psychiatry, Virginia Commonwealth University, Richmond, Virginia, USA; Department of Psychiatry, University of California San Diego, La Jolla, CA, USA; Department of Radiology, University of California San Diego, La Jolla, CA, USA; Program in Neurobehavioral Genetics, Semel Institute, David Geffen School of Medicine, University of California Los Angeles, Los Angeles, CA, USA; Section of Genetic Medicine, University of Chicago, Chicago, Illinois, USA; Neurogenomics Division, The Translational Genomics Research Institute (TGEN), Phoenix, AZ, USA

## Abstract

Attention deficit hyperactivity disorder (ADHD) is a complex disorder with heterogeneous clinical presentations that manifest variability in long-term outcomes. The genetic contributions to this clinical heterogeneity, however, are not well understood. Here, we study 14 084 individuals diagnosed with ADHD to identify several genetic factors underlying clinical heterogeneity. One genome-wide significant locus was specifically associated with an autism spectrum disorder (ASD) diagnosis among individuals diagnosed with ADHD and it was not previously associated with ASD nor ADHD, individually. We used a novel approach to compare profiles of polygenic scores for groups of individuals diagnosed with ADHD and uncovered robust evidence that biology is an important factor in on-going clinical debates. Specifically, individuals diagnosed with ASD and ADHD, substance use disorder (SUD) and ADHD, or first diagnosed with ADHD in adulthood had different profiles of polygenic scores for ADHD and multiple other psychiatric, cognitive, and socio-behavioral traits. A polygene overlap between an ASD diagnosis in ADHD and cognitive performance was replicated in an independent, typically developing cohort. Our unique approach uncovered evidence of genetic heterogeneity in a widely studied complex disorder, allowing for timely contributions to the understanding of ADHD etiology and providing a model for similar studies of other disorders.

## Introduction

Attention deficit hyperactivity disorder (ADHD) is a multifactorial neurodevelopmental disorder with typical symptom onset in childhood, often persisting into adulthood, and affecting many aspects of life through impaired attention, impulsivity and hyperactivity^1,2^. Clinical presentations are heterogeneous and diagnosed individuals demonstrate substantial variability in symptom severity, predominance and duration, treatment, age at time of diagnosis, need for hospitalization, and other long-term outcomes^3-5^. Some diagnosed individuals experience more negative outcomes such as criminality^6^, premature mortality^7^, poor educational attainment^8^, or lower socioeconomic status^9^. Both psychiatric and somatic comorbidities in ADHD are common but their prevalence and (co)occurrence can vary substantially among individuals and across the life-span^3,10^. Better understanding the etiology of clinical heterogeneity is important for improving long term outcomes and precision care. Although genetic heterogeneity is proposed as a feature of complex disorders^11^ and important to understand^12^, insights have proven elusive, especially when considering the extreme polygenic architectures of psychiatric disorders (e.g., ^13,14^).

Family studies show that genetic factors play an important role in ADHD etiology, with estimates of the narrow-sense heritability (*h*^*2*^) around 0.74^15^. This genetic contribution is complex, including copy number variants (CNVs)^16,17^, rare protein truncating variants (PTV)^18^, and polygenes - the numerous common variants with small, independent, additive effects on liability^19^. Estimates of SNP-based heritability (*h*^*2*^_*SNP*_) for ADHD imply this polygene contribution is substantial (22%)^19^. ADHD associated polygenes are known to be pleiotropic, shared broadly across several clustered psychiatric, cognitive, and socio-behavioral traits, and concentrated in chromatin with active roles in neural development and functioning^19,20^. These insights come predominantly from consortia-driven meta-analyses^21,22^ that aggregate multiple cohorts sampled according to different ascertainment criteria, case-control definitions, and healthcare contexts. Mapping robustly associated individual loci via genome wide association studies (GWAS) does not appear overly sensitive to such differences in contributing cohorts, however, inferences from polygenes may be more susceptible^23-25^. Thus, a polygene contribution to clinical heterogeneity in ADHD is plausible and has important implications for study design, across cohort replication, and providing a more nuanced picture of ADHD etiology^12^.

Recent studies have sought robust evidence of genetic heterogeneity for numerous complex disorders^12^, but many have been limited by conceptual, methodological, and data-related challenges that are exemplified by, but not limited to, ADHD. When considering ADHD, few cohorts have been ascertained with both the requisite statistical power for genetic analysis and the necessary depth and breadth of adjacent phenotyping for systematic investigations of clinical heterogeneity. As a result, analyses targeting such heterogeneity are often conducted *post-hoc* or secondary to other aims (i.e., mapping disorder associated loci) where they may also be limited to a focus on single trait or selected variants. This includes, as examples, the association of previously discovered ADHD risk variants with comorbid substance use disorder (SUD)^26^, autism spectrum disorder (ASD)^27^, sex-differences^28^, or symptom persistence^29^. More generally, state of the field analytical approaches (e.g.,^30^) often specifically emphasize variants with an *a priori* association to disorder onset and may miss or under prioritize contributions from modifier variants^31,32^ that could alter clinical trajectories without such a prior association (e.g., those disrupting drug metabolizing enzymes). Clinical presentations of complex disorders vary on a multitude of phenotypic dimensions and likely due to an equally diverse set of genetic factors, so studies that can overcome some of these prior limitations are poised to have wide-reaching impact.

Here, we use data from iPSYCH2012 case-cohort study^33^, which includes the largest single cohort of individuals clinically diagnosed with ADHD (N=14,084) and is linked with a wealth of adjacent phenotyping from Danish population-wide health and civil registers^34-37^. We first define a collection of *ADHD-adjacent traits*, using this term to refer to plausibly relevant, clinical phenotypes of individuals diagnosed with ADHD that we then assess for etiological relevance. We implement a well understood estimator of *h*^*2*^_*SNP*_ in a novel way to prioritize ADHD-adjacent traits that most strongly associate with genetic differences among diagnosed individuals. Then, we conduct GWAS to identify single variants associated with these prioritized traits and describe plausible biological mechanisms. We then introduce a novel polygenic profiling approach that uses multivariate, multinomial logistic regression. This method simultaneously compares healthy controls and multiple case groups across sets of mutually adjusted polygenic scores (PGS) for cognitive, psychiatric, and socio-behavioral traits, while accounting for primary disorder (e.g., ADHD) PGS and covariates. We use this to identify specific and across PGS evidence of genetic heterogeneity. Finally, as an external validation of our findings, we construct PGS using variant effects from ADHD-adjacent trait GWAS to predict cognitive and behavioral performance in an independent, typically developing cohort. Our study adds robust, new genetic perspectives on existing clinical debates surrounding ADHD etiology and can serve as a model for similar investigations in other complex disorders.

## Results

### ADHD-adjacent traits associate with genetic variability among diagnosed individuals

We defined 22 ADHD-adjacent traits from national register data to broadly describe variability in medication use, psychiatric comorbidity, need of care, and sex recorded at birth for 14,084 individuals diagnosed with ADHD (**Methods, Table 1, Supplementary table 1**). We estimated the SNP heritability (*h*^*2*^_*SNP*_) of each trait within the ADHD case group to prioritize those most relevant for follow up with more detailed investigation. (**Methods, Figure 1, Supplementary table 2**). We observed significant *h*^*2*^_*SNP*_ (p<0.002, adjusted for 22 tests) for three traits: a first ADHD diagnosis as an adult (≥18 years of age) (*h*^*2*^_*SNP*_=0.143, s.e.=0.025, p=1.7×10^−10^), an ADHD-adjacent diagnosis of ASD (*h*^*2*^_*SNP*_=0.091, s.e.=0.024, p=2.7×10^−5^), and an ADHD-adjacent diagnosis of SUD (*h*^*2*^_*SNP*_=0.085; s.e.=0.024; p=1.0×10^−4^). An additional seven traits had nominally significant p-values (p<0.05), suggesting with additional samples more traits could be considered for follow up. We used hierarchical clustering to identify patterns in tetrachoric correlations of all pairs of traits. There were roughly three groups: male sex-childhood diagnoses, medication traits, and adult first ADHD diagnosis-mood diagnoses (**Supplementary table 3**, clustered heatmap: **Supplementary figure 1**). We note prioritized traits are at least partially independent with a first ADHD diagnosis as an adult positively associated with an SUD diagnosis (r_TET_=0.62, s.e.=0.01, p<1×10^−10^) and negatively associated with an ASD diagnosis (r_TET_= −0.43, s.e.=0.017, p<1×10^−10^). SUD diagnoses were negatively associated with ASD diagnoses (r_TET_= −0.33, s.e.=0.019, p<1×10^−10^). ADHD-adjacent traits appear widely and robustly associated with genetic differences and further investigations are warranted for a first ADHD diagnosis as an adult, an ADHD-adjacent ASD diagnosis, and an ADHD-adjacent ASD diagnosis.

**Table 1.**
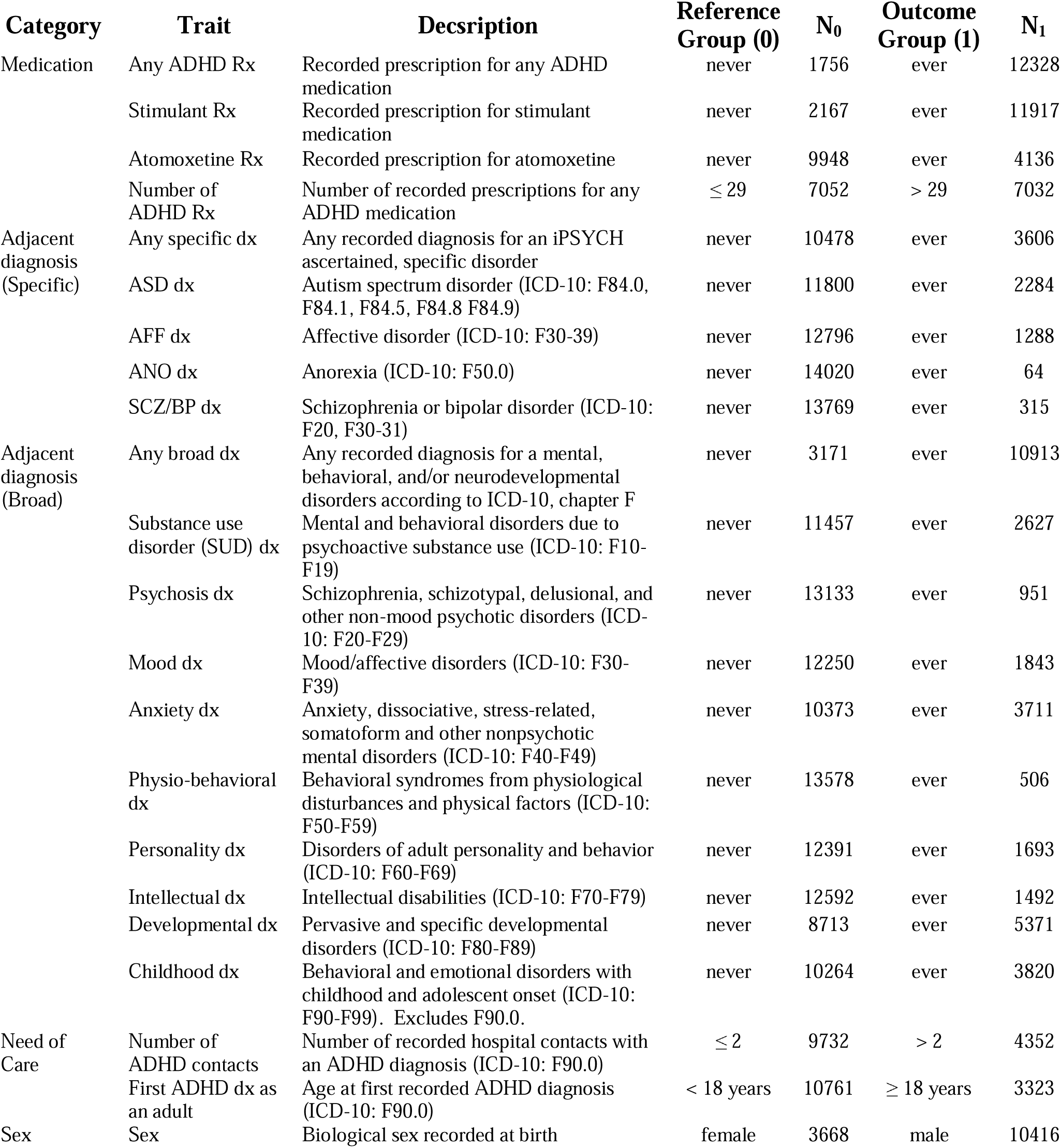
14,084 individuals diagnosed with ADHD vary on a number of clinically relevant adjacent traits. Dx, diagnosis; Rx, prescription; ICD-10, international classiciation of disease, 10^th^ revision; ADHD, attention-deficit hyperactivity disorder.

**Fig. 1.**
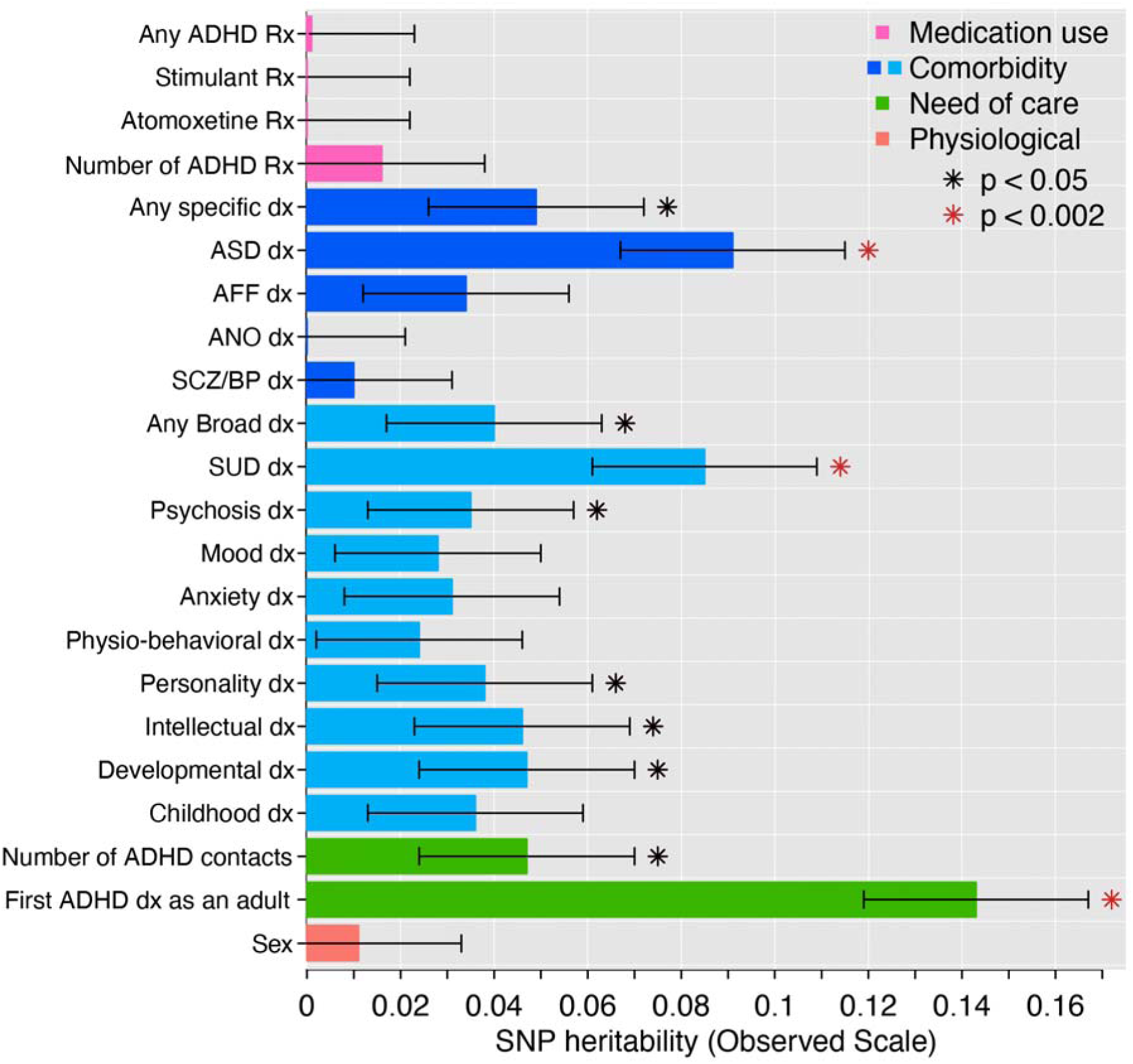
ADHD-adjacent traits associate with genetic variability among diagnosed individuals. The same 14,084 individuals diagnosed with ADHD were repeatedly partitioned into 22 groups on the basis of ADHD-adjacent traits and the SNP-heritability of each trait was estimated with GCTA. Full statistical results are available in Supplementary table 3. Significance (red star) is after Bonferroni correction for 22 tests. Rx, prescription; dx, diagnosis; ADHD, attention-deficit hyperactivity disorder; ASD, autism spectrum disorders; AFF, affective disorders; ANO, anorexia; SCZ/BP, schizophrenia or bipolar disorder; SUD, substance use disorder.

#### rs8178395 is specifically associated with an ADHD-adjacent ASD diagnosis

We performed three GWAS to identify single variants associated with the prioritized ADHD-adjacent traits (**Methods**). No individual SNPs were associated (p>5×10^−8^) with a first ADHD diagnosis as an adult (**Supplementary figure 2**) or an ADHD-adjacent SUD diagnosis (**Supplementary figure 3**). However, one locus on chromosome 17q22 was significantly associated with an ADHD-adjacent ASD diagnosis (hg19: 55,341,733-57,341,733, lead SNP: rs8178395, minor allele (T) frequency (MAF)=0.14, odds ratio (OR)=1.30, s.e. on ln scale = 0.05, p_GWAS_=1.98×10^−08^; **Figure 2a, Supplementary figure 4**). This locus was not identified in previous ADHD^19^ (rs8178395, p=0.44), ASD^38^ (rs8178395, p=0.26), or across-psychiatric disorders^39^ (rs8178289, a proxy SNP with r^2^ LD=0.9, p=0.23) GWAS, although these individuals were included in each meta-analysis. PheWAS for rs8178395 and rs8178289 showed evidence for prior associations with physiological measures of blood cell composition, protein levels, metabolites, cardiovascular complications, and sleep behavior (**Supplementary figure 5, Supplementary table 4**). Quality checks suggest rs8178395 is reliably imputed (**Supplementary figure 6, Supplementary figure 7, Supplementary table 5)**.

**Fig. 2.**
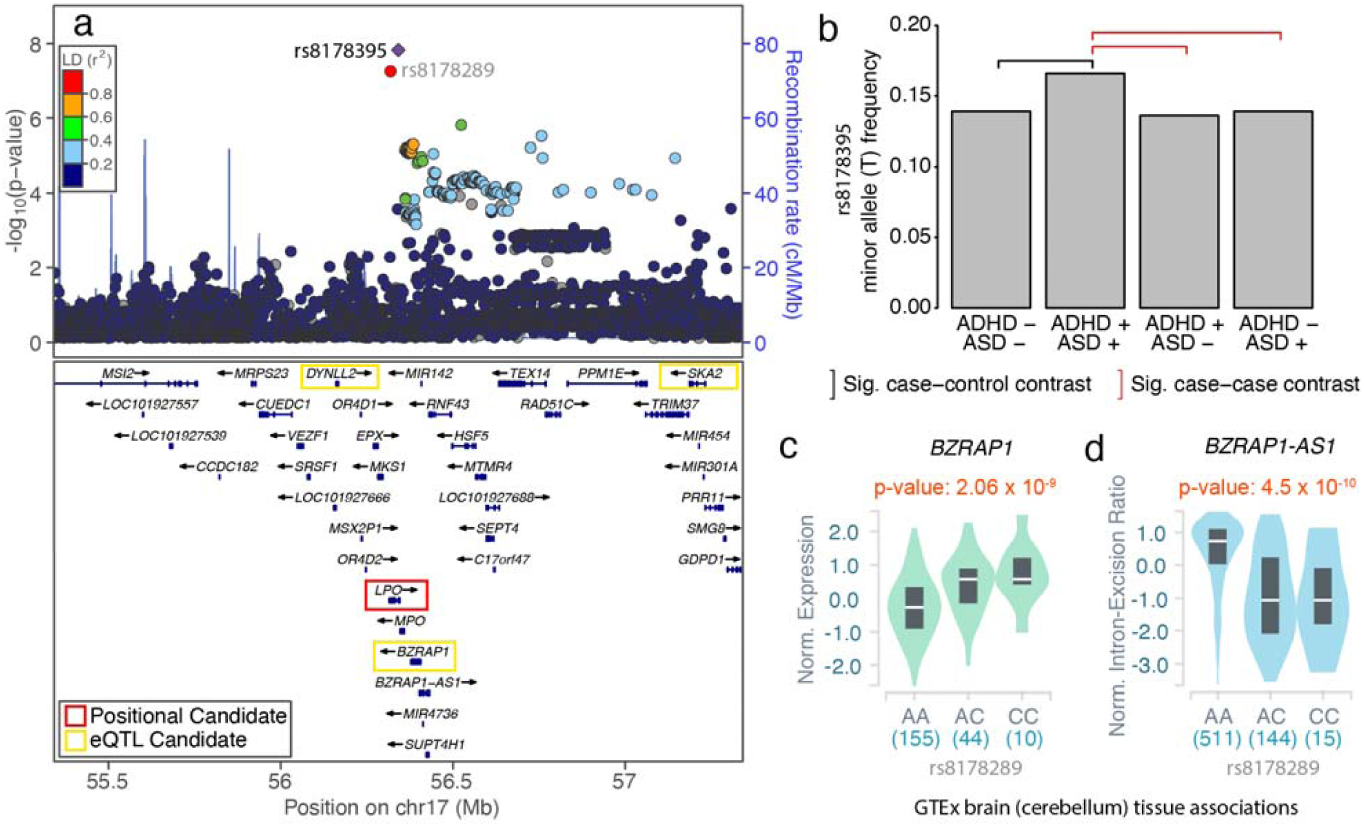
rs8178395 is specifically associated with an ADHD-adjacent ASD diagnosis. **a**, Locus zoom of lead SNP (rs8178395) identified in GWAS comparing individuals diagnosed with both ADHD and ASD to those only diagnosed with ADHD (iPSYCH2012_ASD). *LPO, DYNLL2, BZRAP1*, and *SKA2* are prioritized as candidate genes according to different criteria. LD, linkage disequilibrium; cM, centimorgans; Mb, megabases; TAS, transcription association study; **b**, The minor allele (T) of rs8178395 is significantly increased in frequency in the group of individuals diagnosed with both ADHD and ASD (ADHD +, ASD +) relative to those diagnosed with neither ADHD, nor ASD (black bracket; ADHD -, ASD -: p=2.9×10^−7^) and either, exclusively (red brackets; ADHD +, ASD -: p=2.4×10^−8^; ADHD -, ASD +: p=1.0×10^−6^). Significance is based on multinomial logistic regression. See Supplementary table 4 for additional details. Sig, Significant. **c**, The proxy SNP for rs8178395 (rs8178289, r^2^ LD: 0.9) is associated with *BZRAP1* expression in brain (and other) tissue(s) in GTEx. **d**, rs8178289 is also reported in GTEx as a member of a haplotype containing a splice QTL for a *BZRAP1* associated antisense RNA, *BZRAP-AS1* in the same (and other) tissue(s).

To determine if this variant is a novel ASD SNP or represents evidence of genetic heterogeneity within ADHD, we employed multinomial logistic regression (MLR) (**Methods**). This test extends the two-group logistic regression comparing ADHD with or without an adjacent ASD diagnosis to additionally compare individuals with an ASD, but not ADHD, diagnosis and controls with neither diagnosis. We observed that the frequency of the minor (T) allele of rs8178395 was significantly increased in the group with ADHD-adjacent ASD relative to all other groups (vs. ADHD only, p_MLR_=2.4×10^−8^; vs. ASD only, p_MLR_=1.0×10^−6^; vs. controls, p_MLR_=2.9×10^−7^; **Figure 2b, Supplementary table 6)**. rs8178395 indexes the first locus reported to have a specific association to individuals diagnosed with both ADHD and ASD.

The locus indexed by rs8178395 contains 38 genes (**Figure 2a**). rs8178395 falls within an intron of *LPO* and has been associated with expression of *DYNLL2, BZRAP1*, and *SKA2* in brain and *MPO, RAD51C, RNF43, SEPT4, SMG8, SUPT4H1, TEX14*, and *TRIM37* in other tissues (**Methods, Supplementary table 7)**. Partitioned LD-score regression did not identify significant enrichment in any selected adult or fetal brain derived genome annotations (**Methods, Supplementary figures 8-10, Supplementary table 8**). So, we performed a regional transcription association study (TWAS) (**Methods**) using expression levels imputed via multiple adult^40^ and fetal brain^41^ expression quantitative trait loci (eQTLs) (**Methods**; **Supplementary table 9**). This did not identify significant candidates after Bonferroni correction (p<0.006); imputed adult-brain *SKA2* expression was the most significant (p=0.008). The proxy SNP for rs8178395 (rs8178289, r^2^ LD=0.9) is also associated with *BZRAP1* expression in brain tissue (**Figure 2c**) and, additionally, is reported by GTEx as a member of a haplotype containing splice QTLs (sQTL) for an associated antisense RNA, *BZRAP-AS1*, active in brain (**Figure 2d**) and other tissues^42^. Regulation of *BZRAP1* may be an especially plausible mechanism for this association. *BZRAP1* is a brain expressed gene^43^ encoding a binding protein that couples voltage gated calcium channels to neurotransmitter vesicles in the presynaptic active zone^44^, has a well-characterized role in neurotransmitter release and synaptic transmission^45^, and exonic CNVs have been associated with ASD in multiplex families^46^.

### ADHD-adjacent traits share polygenes with psychiatric, cognitive, and socio-behavioral traits

Next, we pursued a novel approach for mapping *poly*genetic contributions to clinical heterogeneity, extending concepts used in studies of disorder onset to increase power beyond single locus tests. We first estimated the genetic correlations (ρ_G,SNP_; **Methods**) between ADHD-adjacent traits and 41 psychiatric, cognitive, and socio-behavioral traits using GWAS summary statistics (**Figure 3, Supplementary table 10**). Trends were broadly similar across reference traits for a first ADHD diagnosis as an adult and ADHD-adjacent SUD, and generally opposite to those estimated for ADHD-adjacent ASD, consistent with phenotypic correlations **(Supplementary figure 1, Supplementary table 3)**. Individually, an adult first diagnosis was genetically correlated (ρ_G,SNP,_ FDR<0.05) with psychiatric traits, reproductive behaviors, education, and cognitive performance (**Figure 3a**), and, of note, showed a large negative trend with clinically ascertained and defined childhood ADHD^47^ (ρ_G,SNP_= −0.5, s.e.=0.27, p=0.07). ADHD-adjacent SUD showed similar estimates of ρ_G,SNP_ with psychiatric outcomes, reproductive behaviors, education, and cognitive performance, but also smoking behavior (**Figure 3b**). ADHD-adjacent ASD, however, had different ρ_G,SNP_ for education and cognitive performance (**Figure 3c**). In summary, ADHD-adjacent traits likely share polygenes with psychiatric, cognitive, and socio-behavioral traits and may define genetic heterogeneity in ADHD.

**Fig. 3.**
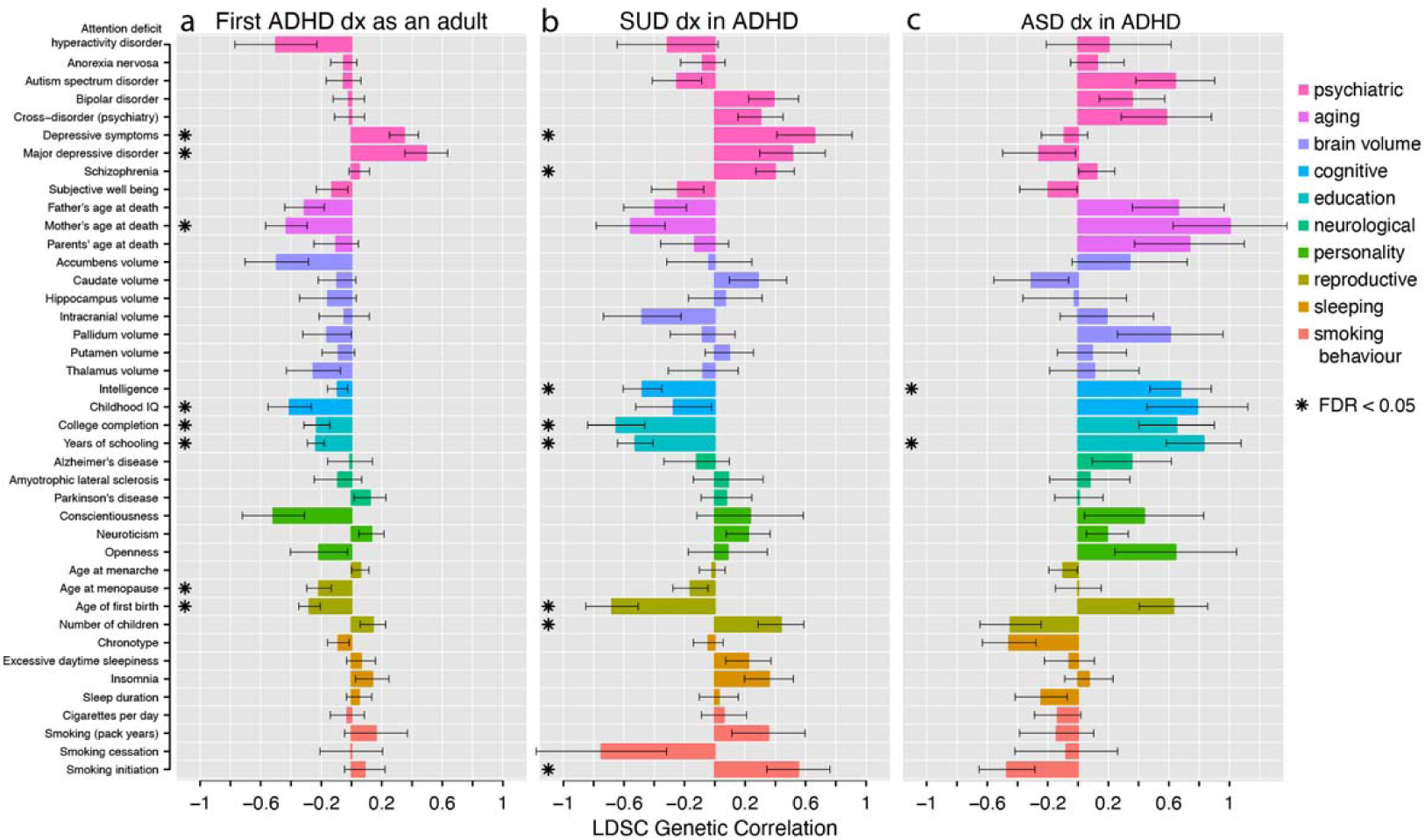
ADHD-adjacent traits share polygenes with psychiatric, cognitive, and socio-behavioral traits. LD score regression was used to estimate the genetic correlations between ADHD-adjacent traits and a reference set of psychiatric, cognitive, and socio-behavioral traits. **a**, A first diagnosis of ADHD as an adult, **b**, an ADHD-adjacent SUD diagnosis, and **c**, an ADHD-adjacent ASD diagnosis show different patterns of genetic correlation with 41 reference traits. Prioritizationd determined by FDR < 0.05. Error bars denote standard error of estimates. See Supplementary table 10 for full statistical results. LDSC, LD score regression; FDR, false discovery rate; dx, diagnosis; ADHD, attention-deficit hyperactivity disorder; ASD, autism spectrum disorders; SUD, substance use disorder.

### Polygenic scores for psychiatric, cognitive, and socio-behavioral traits define aspects of heterogeneity in ADHD

We extend these more qualitative descriptions with a novel polygenic profiling approach providing robust, quantitative tests that ADHD-adjacent traits manifest as a result of genetic heterogeneity indexed by profiles of PGS (**Methods**; **Figure 4, Supplementary tables 11-13**). The multi-PGS profiles were different among individuals receiving a first diagnosis for ADHD as an adult (1^st^ Dx adult; n=3,323), a first diagnosis for ADHD before adulthood (1^st^ Dx child; n=10,761), and controls (ADHD-; n=21,409). The mean PGS for ADHD, depressive symptoms, childhood IQ, years of schooling, and age at first birth varied significantly among the three groups (p<4×10^−4^; **Figure 4a**, stars; **Supplementary table 11**) and these three-group-wise differences were driven by specific, highly significant pairwise contrasts (**Figure 4a**, black and red brackets). The two ADHD case groups generally deviated significantly from controls and in the same direction. Individuals first diagnosed as an adult had, on average, significantly less positive PGS (i.e., closer to the population average) for ADHD and more negative PGS (i.e., further from the population average) for years of schooling than ADHD cases diagnosed before adulthood. Additional trends suggested more negative cognitive, education, and reproductive PGS and more positive psychiatric PGS for adult diagnosed ADHD cases. Simulations suggest the overall trends here are not consistent with models where an adult diagnosis is simply due to ADHD liability associating with onset, a misdiagnosis of MDD, or education as an age dependent, heritable exposure (**Supplementary Note, Supplementary Figures 11-15**).

**Fig. 4.**
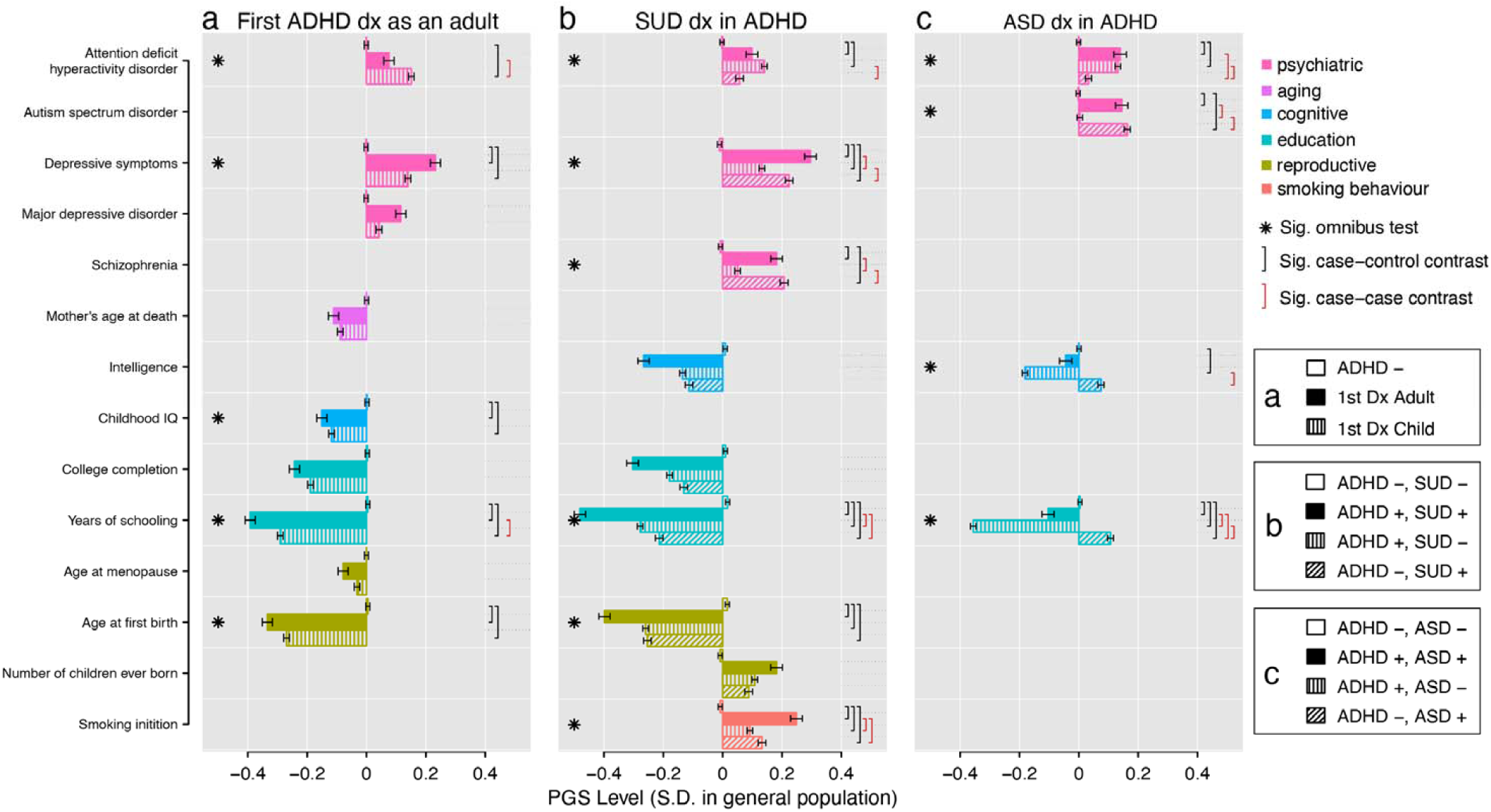
Profiles of polygenic scores for psychiatric, cognitive, and socio-behavioral traits define aspects of heterogeneity in ADHD. The mean level of polygenic scores (PGS) are displayed for ADHD subgroups, controls, and complementary disorder groups after centering and standardizing based on a population random sample. **a**. Individuals not diagnosed with ADHD (ADHD -), first diagnosed as an adult (1^st^ Dx Adult), and first diagnosed as a child (1^st^ Dx Child), **b**. individuals diagnosed with neither ADHD, nor SUD (ADHD-, SUD-), both ADHD and SUD (ADHD+, SUD+), ADHD but not SUD (ADHD+, SUD-), and SUD but not ADHD (ADHD-, SUD+), and **c**. individuals diagnosed with neither ADHD, nor ASD (ADHD-, ASD-), both ADHD and ASD (ADHD+, ASD+), ADHD but not ASD (ADHD+, ASD-), and ASD but not ADHD (ADHD-, ASD+), vary significantly with respect to multivariate profiles of PGS. Stars denote a significant global test for differences in PGS level among all groups, while brackets denote significant pairwise group contrasts, accounting for all other PGS and 25 ancestry principle components in a joint multinomial logistic regression. Bonferroni adjustment for 127 contrasts. Errors bars denote standard error of mean. See Supplementary tables 11-13 for complete statistical results. ADHD, attention deficit hyperactivity disorder; ASD, autism spectrum disorder; SUD, substance use disorder; Dx, diagnosis; Sig., Significant.

The multi-PGS profiles for individuals with an ADHD-adjacent SUD diagnosis (ADHD+, SUD+; n=2 627), without an ADHD-adjacent SUD diagnosis (ADHD+, SUD-; n=11 457), with an SUD but no ADHD diagnosis (ADHD-, SUD+; n=5 943), and with neither ADHD nor SUD diagnoses (ADHD-, SUD-, controls; n=20 509) were also different and with a similar pattern to the previous result (**Figure 4**). The mean PGS for ADHD, depressive symptoms, schizophrenia, education as in years of schooling, age at first birth, and smoking initiation varied significantly among the four groups (p<4×10^−4^; **Figure 4b**, stars; **Supplementary table 12**). As above, all case groups generally deviated from controls significantly and in the same direction. Notable pairwise differences (**Figure 4b**, black and red brackets) included individuals with ADHD-adjacent SUD diagnoses showing a trend towards less positive ADHD PGS, significantly more positive PGS for depressive symptoms, schizophrenia, and smoking initiation, and significantly more negative PGS for years of schooling, that ADHD without an adjacent SUD diagnosis. Individuals with ADHD-adjacent SUD diagnoses showed, relative to those with SUD diagnoses only (i.e., no ADHD diagnosis), significantly more negative PGS for education and more positive PGS for smoking initiation, but no significant differences in ADHD, depressive symptoms, or schizophrenia PGS.

The multi-PGS profiles for individuals with ADHD-adjacent ASD diagnoses (ADHD+, ASD+; n=2 284), with ADHD but no adjacent ASD diagnosis (ADHD+, ASD-; n=11 800), with ASD but not ADHD diagnoses (ADHD-, ASD+; n=9 804), and with neither (ADHD-, ASD-controls; n=21 197) were also different, but trends were broadly different from the previous two analyses. The mean PGS for ADHD, ASD, intelligence, and years of schooling varied significantly among the four groups (p<4×10^−4^; **Figure 4c**, stars, **Supplementary table 13**). The individuals with ADHD-adjacent ASD diagnoses had profiles that appeared to reflect both single-diagnosed groups, simultaneously. The ADHD-adjacent ASD group had significantly more positive ADHD and ASD PGS than controls, significantly more positive ASD PGS than, but similar ADHD PGS as, the ADHD only group, and significantly more positive ADHD PGS than, but similar ASD PGS as, the ASD only (**Figure 4b**, black and red brackets). Interestingly, the single diagnosed ASD and ADHD groups had opposite trends (i.e., above vs. below the population average) for intelligence and education PGS, and the ADHD-adjacent ASD group fell nearly perfectly in between the two. Simulations suggest the overall trends here are not consistent with misdiagnosis and most consistent with models where diagnosed individuals have diagnosis that relate to an excess of liability for both disorders, simultaneously (**Supplementary Note, Supplementary Figures 16-24**).

Large differences among groups that are not described as statistically significant (e.g., **Figure 4b:** college completion) are due to collinearity among constituent profile PGS that are fitted jointly (**Methods**) and are significant when fitted separately (**Supplementary figure 25, 26, Supplementary tables 14-16**). Trends depicted as unadjusted levels of PGS in **Figure 4** are directionally consistent with trends when depicted as partial PGS effects (i.e., regression coefficients) estimated in the joint models (**Supplementary figure 27, Supplementary tables 11-13**).

### A polygenic score for ADHD-adjacent ASD is associated with cognitive performance in an independent, typically developing cohort

Finally, we sought external support for our finding that polygenic contributions to clinical heterogeneity in ADHD, as indexed by ADHD-adjacent traits, are shared with psychiatric, cognitive, socio-behavioral traits. To pursue this, we used the results of our three ADHD-adjacent trait GWAS to construct PGS in the Adolescent Brain and Cognitive Development (ABCD) study^48^, an independent, typically developing child cohort. Each PGS was tested for an association with each of 51 behavioral and cognitive assessments, adjusting for ADHD PGS and other potential confounders (**Methods, Figure 5, Supplementary table 17**). These analyses confirmed an association between the polygenic basis of an ADHD-adjacent ASD diagnoses and cognitive performance. Specifically, a higher PGS was significantly (p<0.001) associated with increased performance on an assessment of overall cognitive function that appears driven by higher scores in verbal cognition (e.g., crystalized composite, and picture vocabulary scores; **Figure 5c**). No other tests were strictly significant, although a few relevant trends are noted: the signs of the relationships of the different PGS and cognitive or psychiatric traits were in line with our previous analyses and multiple individual PGS trends were consistent for specific psychiatric or behavioral symptoms. We find consistent, independent support for a polygenic relationship between an ADHD-adjacent ASD diagnosis and cognitive performance, but the small discovery GWAS and indirect, modest replication sample limit our power for broader replication.

**Fig. 5.**
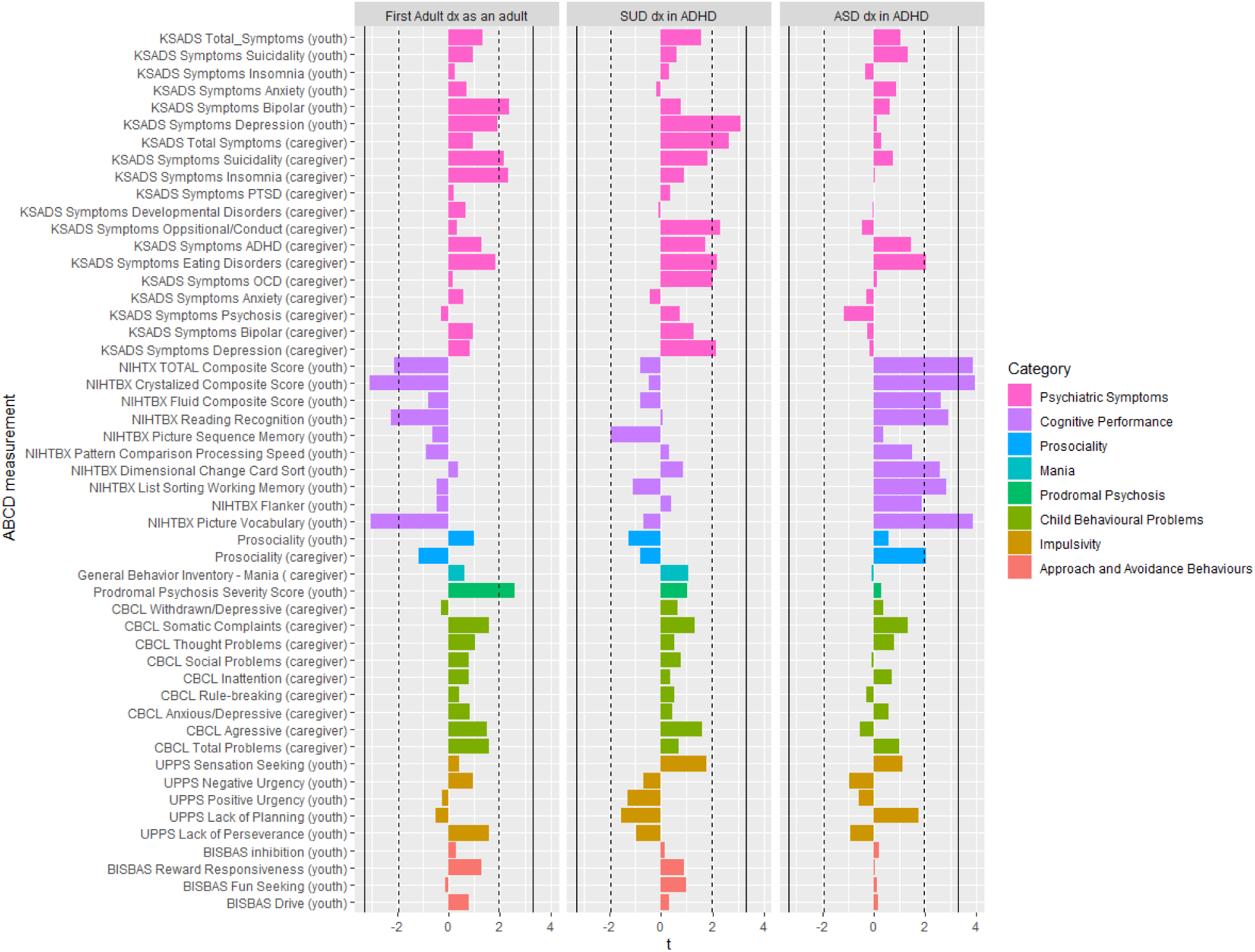
Polygenes for ADHD-adjacent ASD are associated with cognitive performance in an independent, typically developing cohort. Polygenic scores (PGS) constructed using summary statistics from ADHD-adjacent trait GWAS were tested for association with cognitive, behavioral, and psychiatric traits in 5 449 nine or ten year-old children from the Adolescent brain cognitive development (ABCD) Study. No significant associations were found for **a** PGS for adult diagnosed ADHD or **b** ADHD-adjacent SUD, although consistent trends are prevalent. **c** PGS for an ADHD-adjacent ASD diagnosis associated significantly with cognitive performance, replicating our previous finding that polygenes are shared among these two domains. Significance was after Bonferroni correction for 51 tests (solid line) and suggestive trends are noted for p< 0.05 (dotted line). Caregiver or youth in parenthesis denotes the informant for the assessment of the child. t, t-statistic. See Supplementary table 17 for complete statistical results and supplementary table 19 for a description of the questionnaire used for each ABCD assessment.

## Discussion

In this study we used a unique data resource and a novel analysis strategy to identify new evidence for genetic heterogeneity underlying clinical heterogeneity in a widely studied complex disorder. We showed that multiple, clinically relevant ADHD-adjacent traits were related to genetic variability among diagnosed individuals and that these genetic contributions were complex (i.e., marked by few large effects despite significant *h*^*2*^_*SNP*_) but characterizable. Our multivariate analysis of polygenic profiles demonstrated that variability in polygenes associated with psychiatry, cognitive, and socio-behavioral traits is likely to underlie differences in clinical presentations of individuals diagnosed with ADHD. Our findings regarding age of first ADHD diagnosis and ADHD-adjacent ASD diagnoses, in particular, make timely contributions to the understanding of the genetic etiology of ADHD, where other results reflect important themes that are relevant for the study of complex disorders more broadly. Finally, our approach may enable better characterizations of heterogeneity in other complex disorders.

Despite evidence of clinical co-occurrence in individuals and families, shared genetic risk factors^18,49,50^, and biological overlap^51^, no previous GWAS has focused specifically on individuals with diagnoses for both ADHD and ASD^50,52,53^. We identified a locus that appears novel for both disorders, and specific to this doubly diagnosed group. eQTL and previous trait associations suggest the locus is functional, has a plausible connection to regulating neurobiology^43-45^, and the association is supported in a complementary design using multiplex ASD families^46^. While this association needs further replication, it suggests that studying specific comorbidity patterns or presentations may help identify pleiotropic SNPs and disentangle antagonistic or heterogeneous variant effects.

Until the introduction of DSM-5 in 2013^1^, formal guidelines did not allow ASD and ADHD to be diagnosed as comorbid^54^. The recognition of overlap in cognition^55^, genetics^55^, symptoms scores^56^, and neuroanatomic charcteristics^57^ challenge this and the possibility that the core etiologies of both disorders could exist in an individual, simultaneously, are active areas of research and debate^54^. Our polygenic profile approach suggests that individuals with an ADHD-adjacent ASD diagnosis carry, at minimum, the *polygenic* etiologies of both individual disorders. This idea is supported by a recent register study in the Swedish population^50^ that reported comorbid ADHD and ASD diagnosis were most often made on the same day, by the same clinician, observing concurrent symptoms. This trend stands in contrast to similar studies of bipolar disorder and schizophrenia, that suggest an appreciable level of lifetime comorbidity may relate to evolving symptoms and difficulties in diagnosing first episode psychosis^58^. Our polygenic profiling approach supports the notion that ASD and ADHD can be experienced simultaneously, consistent with emerging trends in clinical epidemiology and evolving psychiatry nosology.

The nature of adult-onset ADHD, it’s validity, existence, and whether it represents a distinct clinical or biological construct, is a particularly active and contentious research topic^3,59,60^. The crux of this debate rests on the timing of symptom onset. As above, it was DSM-5^1^ that introduced criteria for an adult ADHD diagnosis. Here, adult ADHD requires retrospective self or informant reported childhood symptoms for a diagnosis, but these reports may suffer recall bias, and objective, representative longitudinal studies of premorbid symptoms are sparse. One perspective (e.g.^29^), then, has been that a first diagnosis as an adult should be thought of as persistent ADHD, perhaps missed during childhood, and, as symptoms may remit with age, be considered a more severe or biological form of ADHD. Here, we observed that, contrary to this perspective, individuals with a first registered diagnosis as an adult had *less* ADHD PGS suggesting they may have a more environmental, less reliable, or less severe form of the disorder^61^. Our polygenic profile approach offers varied support for each of these hypotheses in that adult diagnosed individuals had lower education PGS, higher non-ADHD psychiatric PGS, and similar profiles to individuals with ADHD-adjacent SUD diagnoses. Our data and approach implicate genetic heterogeneity in important debates surrounding adult ADHD where other studies^29^ with similar aims have been less successful.

Beyond ADHD etiology, our study highlights a few themes that are broadly relevant for the study of complex disorders and associated heterogeneity. First, as demonstrated for MDD^23^, male pattern baldness^62^, and in simulations^23,25,63^, the polygenic background of case groups may become skewed if evolving nosology, case definitions, or censoring are not considered carefully. Here, we expect that evolving guidelines around comorbid ADHD and ASD may change the genetic landscape of typical ADHD and ASD case groups, implicitly, and more active ascertainment or censoring mechanisms could have similar consequences. Second, PGS have received a lot of attention as potential clinical instruments for various applications^64,65^, but the focus has often been on primary disorder scores (i.e., here, ADHD) and distinguishing cases from controls. Here we saw using profiles of multiple polygenic scores is a powerful approach for distinguishing among ADHD cases, and that the strongest predictors were not primary disorder PGS. The idea that the polygenic background of a patient can alter a clinical trajectory is not new, but as PGS are aimed at more diverse clinical decisions (i.e., beyond case-control discrimination), more comprehensive summaries of polygenic background should be considered. Finally, methods for detecting heterogeneity might benefit from relaxing strong *a priori* restrictions to onset associated variants while also focusing more broadly on polygenes. Our approach has broad implications for the study of complex disorders beyond ADHD.

Our results must be considered in light of a few important limitations. We use register diagnoses, which are given with high reliability by trained psychiatrists, but are assigned per hospital contact, and may shift over time as patient symptoms evolve or clarify^58^. This could lead to some misdiagnosis. This issue is not unique to register studies as it reflects clinical practice but may offer a different perspective from studies of research diagnoses that may include retrospective censoring or integration according to a hierarchy. Some individuals may seek treatment exclusively through primary care and those contacts are not available in iPSYCH. This may be especially relevant when considering symptom onset and first diagnosis as earlier contacts could be missed. A registered diagnosis may still represent an increase in severity, if not onset, as a hospital referral could represent a change in need of care. The iPSYCH cohort is relatively young and may underestimate the prevalence of adult and later onset outcomes. iPSYCH includes only a combined ADHD subtype (ICD-10: F90.0), and important genetic differences may emerge if individuals diagnosed with inattentive (ICD-10: F98.8) and combined subtypes (ICD-10: F90.1, F90.8, or F90.9) were included. Methodologically, we compared groups of ADHD cases to estimate SNP-heritability and performed GWAS, using these to implicate genetic contributions to clinical heterogeneity of ADHD. However, just as with case-control analysis, these approaches are sensitive to spurious differences. This could occur due to population stratification or if heritable, disorder irrelevant features (e.g., hair color) were used to define patient groups^12,30^. In our work, we mitigate this by adjusting for ancestry related principal components, carefully selecting ADHD adjacent-traits with plausible clinical relevance or specificity to ADHD and including controls in our polygenic profile approach.

This work is part of an emerging body of evidence in psychiatric genetics that suggests we are now, after decades of data aggregation, at a point where we can begin to study not just what makes diagnosed individuals different from healthy controls, but what may differentiate diagnosed individuals from each other with respect to outcomes. Next, we must consider the implications these differences have across multiple areas of research and clinical care.

## Online Methods

### iPSYCH2012 case-cohort study

The Lundbeck Foundation initiative for Integrative Psychiatric Research (iPSYCH)^33^ is a case-cohort study of individuals born in Denmark between 1981 and 2005 (n=1 472 762). 87 764 individuals were sampled, including a random sample of 30 000 and 59 996 ascertained for diagnoses of attention-deficit hyperactivity disorder (ADHD), autism spectrum disorders (ASD), anorexia (ANO), affective disorder (AFF), bipolar disorder (BP), or schizophrenia (SCZ). DNA was extracted from dried bloodspots in the Danish Neonatal Screening Biobank^66^. Diagnoses were obtained from the Danish Psychiatric Central Research Register (PCR)^36^ and for anorexia also from the Danish National Patient Register (DNPR)^35^ from 1 year birthday or 10 year birthday of study individuals to December 31, 2012. Linkage across registers uses the Danish Civil Registration System^37^. Diagnoses given by psychiatrists in primary care are not recorded in these registers. This study focused specifically on 14 084 of 18 726 individuals ascertained for ADHD (ICD-10: F90.0), 12 088 of 16 146 ascertained for ASD (ICD-10: F84.0, F84.1, F84.5, F84.8 or F84.9), and 21 197 of 30 000 controls diagnosed with neither ADHD nor ASD. Furthermore, 8 498 individuals with substance use disorders (SUD) (ICD-10: F1) were selected from among the 30 000 population controls and 57 764 psychiatric cases and finally, 20 509 of 30 000 controls with neither ADHD nor SUD. These subsets passed quality control (see below). The use of this data follows standards of the Danish Scientific Ethics Committee, the Danish Health Data Authority, the Danish Data Protection Agency, and the Danish Neonatal Screening Biobank Steering Committee. Data access was via secure portals in accordance with Danish data protection guidelines set by the Danish Data Protection Agency, the Danish Health Data Authority, and Statistics Denmark.

Genotyping was performed on the Infinium PsychChip v1.0 array with amplified DNA extracted from dried bloodspots. Data quality control is described in detail elsewhere^20,33^. Briefly, 246 369 of the ∼550 000 genotyped SNPs were deemed good quality, phased using SHAPEIT3^67^, and imputed using the 1 000 genomes project phase3^68^ as a reference with Impute2^69^. Imputed additive genotype dosages and best-guess genotypes were checked for imputation quality (INFO>0.2), Hardy–Weinberg equilibrium (HWE; p <1×10^−6^), association with genotyping wave (p<5×10^−8^), association with imputation batch (p<5×10^−8^), differing imputation quality between cases and controls (p<1×10^−6^), and minor allele frequency (MAF>0.01). 8 019 760 dosages and best-guess genotypes remained. Subjects of homogeneous genetic ancestry were selected after principal components analysis using EIGENSOFT v6.0.1^70^. One from each pair of individuals with closer than third degree kinship as estimated with KING v1.9^71^ was excluded, and no samples had abnormal heterozygosity, high levels of missing genotypes (>1%), nor genotype/recorded sex discordance.

#### Selecting ADHD-adjacent traits

DNPR^35^ and PCR^36^ contain information (e.g., date of admission, ICD diagnostic code, etc.) on all inpatient hospital contacts since 1977 and 1969, respectively, and outpatient and emergency room contacts since 1995. From these registers we defined two sets of ADHD-adjacent traits to capture psychiatric comorbidity. First, we coded the presence of specific psychiatric disorders as defined by the iPSYCH2012 case-cohort ascertainment criteria^33^, including ASD, AFF, ANO, and combined psychotic disorders (SCZ and BP). As a second set, we summarized psychiatric diagnoses more broadly, recording each ICD-10 Behavioral and Mental Disorders subchapter^2^ recorded in either PCR or DNPR until 2016 (F1, F2, F3, F4, F5, F6, F7, F8, F9 excluding ADHD (F90.0-9 and F98.8)). As before, we created variables representing a diagnosis from each subchapter. We also counted the number of hospitalizations with ADHD (ICD-10 F90.0) recorded as the main diagnosis of action in the PCR up until 2016, also dichotomized as a split by the median. A variable splitting cases on the first recorded ADHD diagnosis (ICD-10 F90.0) occurring before or after 18 years of age. Finally, a variable was created recording sex as reported at birth.

The Danish National Prescription Register (NPR)^34^ holds information on prescriptions redeemed from pharmacies in Denmark since 1995. The NPR does not cover drugs used during hospital admissions, by certain institutionalized individuals (e.g. psychiatric), or drugs supplied directly by hospitals or treatment centers. We defined three traits from the NPR noting cases with a record of at least two prescriptions after the age of 3 for drugs with the Anatomical Therapeutic Chemical (ATC) codes for stimulants (N06BA01, N06BA12, N06BA02, N06BA04), or atomoxetine (N06BA09), or both (i.e., any ADHD medication). A fourth trait counted the number of prescriptions an individual had filled and dichotomized on the median number (in this cohort) of total prescriptions. Further details are provided in **Supplementary table 1**.

We used tetrachoric correlations to describe the co-occurrence of ADHD-adjacent traits, estimating them with the R package *polycorr*^*72*^ and adding 0.5 to zero cells^73^. Hierarchical clustering was performed using the *heatmaply*^*74*^ package.

#### SNP-based heritability

SNP heritability (*h*^*2*^_*SNP*_) was estimated on the observed scale using GREML in GCTA v1.92.1 beta6^75^ with 25 ancestry principal components as fixed effect covariates. The genetic relationship matrix (GRM) was constructed using GCTA and from best-guess genotypes with a MAF greater than 0.01. Significance was adjusted by Bonferroni correction (p<0.05/22=0.002), and nominally significant tests (p<0.05) were deemed suggestive.

#### Genome-wide Association Studies

Case-case genome-wide association studies (GWAS) were performed within the ADHD case group (n=14 084). Logistic regression in plink v1.90b3.34^76^ was used to test the association between imputed additive allele counts and case subgroup membership, with 25 ancestry principal components as covariates. Genomic inflation was estimated as the unconstrained LD-score regression^77^ intercept following: https://github.com/bulik/ldsc/wiki. Individual SNPs with p<5×10^−8^ were declared genome-wide significant. Lead SNPs were defined as the most significant SNP within a 2 mega base (mb) locus.

For genome-wide significant loci, multinomial logistic regression (MLR)^78^ was performed using the R package *nnet*^79^. The MLR tests the logistic regression comparison of allele counts at a lead SNP between ADHD subgroups (i.e., individuals with comorbid ADHD and ASD, n=2 284, and individuals with ADHD but not ASD, n=11 800) to simultaneously include comparisons with individuals diagnosed with relevant complementary disorder (i.e., individuals with ASD but not ADHD, n=9 804) and undiagnosed controls (i.e., individuals with neither ADHD nor ASD, n=21 197).

For genome wide significant loci, candidate genes were selected according to three criteria: 1) positional candidate genes were selected by ANNOVAR^80^ in FUMA^81^ as overlapping a lead SNP’s genomic position, 2) eQTL candidate genes with previous expression association to a lead SNP in one of multiple sources aggregated by FUMA, 3) transcriptional association candidate genes with predicted expression in adult and/or fetal brain. Here a transcription association (TWAS) analysis was used integrating published per-SNP effects on expression for fetal^41^ and adult^82^ brain, and FUSION^83^ to generate aggregate predicted expression in each individual. TWAS p-values were considered significant after Bonferroni correction (p<0.05/9=0.006). For lead SNPs from genome-wide significant loci, the GWASatlas v20191115^84^ (https://atlas.ctglab.nl/) was used to pursue a pheWAS across 4 756 studies grouped according to the provided ontology. The proxy SNP for rs8178395 was identified using LDlink (https://ldlink.nci.nih.gov/?tab=help#LDproxy)^85^.

We used LD score regression (LDSC)^77^ to partition *h*^*2*^_*SNP*_ among selected genome annotations, accounting for LD and baseline annotations. Annotations of interest included those defined by eQTLs from fetal brain^41^, eQTLs from adult^40^ brain, and regions of open chromatin measured by ATAC sequencing in fetal^86^ and adult^87^ brain. A “full baseline model” of 53 functional categories was employed following Finucance et al^88^. We followed the author protocols for analysis (https://github.com/bulik/ldsc/wiki/Partitioned-Heritability). eQTL and ATAC LD-scores were created for the subset of human SNPs genotyped in HapMap v3 SNPs (HM3) using the LD-score regression software by identifying HM3 SNPs within a 500bp window (±250) around each eQTL SNP or within an ATAC open chromatin window. Category-specific LD-scores were the sum of LD (r^2^) for each HM3 SNP with SNPs meeting the previous functional criteria. The European subset of the 1000 Genomes Project Phase3 was used as an LD reference. Enrichment was estimated for each annotation as the proportion of heritability explained by each annotation divided by the proportion of SNPs in the genome falling in that category. Enrichment p-values were declared significant after Bonferroni correction (p<0.05/28=0.0018) or suggestive when p<0.05.

#### SNP-based genetic correlations

SNP-based genetic correlations (ρ_G,SNP_) were estimated using LD-Score regression in LD-hub (http://ldsc.broadinstitute.org/)^89^, an online-tool for estimating ρ_G,SNP_ against a catalog of published GWAS studies. Summary statistics were uploaded for the three ADHD variable case-case GWAS performed for this study and were estimated with each of 41 cataloged traits from 10 psychiatric, cognitive, and socio-behavioral categories: smoking, neurology, personality, sleeping, cognition, reproduction, education, neuroimaging, psychiatry, and ageing. Prioritized traits had FDR<0.05.

#### Polygenic Scores in iPSYCH

GWAS summary statistics (**Supplementary table 18**) were downloaded from public repositories and PGS for each were calculated using LDpred v01 for individuals in iPSYCH^90^. Reference GWAS were selected to ensure no subject overlap with iPSYCH. SNP inclusion criteria were: MAF>0.05, INFO>0.98, and HWE p-value>1×10^−5^. Palindromic (A/T, C/G) SNPs, SNPs not uniquely mapped to hg19 positions, and SNPs not having unique IDs in dbSNP v151 were excluded. Only one SNP from a group of SNPs in LD (pairwise r^2^ LD>0.99) was retained. We used an LDpred p-parameter of 1, corresponding to an infinitesimal model as a prior assumption for the per-SNP effects.

We used multivariate, multinomial logistic regression (MLR)^78^ implemented in the R package *nnet*^*79*^ to test for heterogeneity in PGS profiles among ADHD case subgroups, non-diagnosed controls, and individuals diagnosed with ASD or SUD but not ADHD. For each of the three ADHD variables, we jointly fit primary disorder PGS (i.e., ADHD, ASD), the set of PGS for traits that showed significant ρ_G,SNP_ with the ADHD variable in the LDSC analysis described above, and 25 ancestry principal components. MLR models fitting each PGS individually, along with ancestry covariates, are presented in the **Supplementary tables 14-16**. Three MLR were of primary interest. First, for ADHD-adjacent ASD, we compared individuals diagnosed with neither ADHD nor ASD (n=21 197), both ADHD and ASD (n=2 284), ADHD but not ASD (n=11 800), and ASD but not ADHD (n=9 804) on PGS profiles containing scores for ADHD^47^, ASD^91^, intelligence^92^ and years of schooling^93^. For ADHD-adjacent SUD, we compared individuals with neither ADHD nor SUD (n=20 509), both ADHD and SUD (n=2 627), ADHD but not SUD (n=11 457), and SUD but not ADHD (n=5 943) on PGS profiles containing scores for ADHD^47^, depressive symptoms^94^, schizophrenia^95^, intelligence^92^, college completion^96^, years of schooling^93^, age at first birth^97^, number of children ever born^97^, and smoking initiation^98^. For age of first ADHD diagnosis we compared individuals without a diagnosis for ADHD (n=21 409), with a first diagnosis of ADHD as an adult (>18 years of age) n=3 323), and with a first diagnosis of ADHD before age 18 (n=10 761) on PGS profiles containing scores for ADHD^47^, depressive symptoms^94^, major depressive disorder^99^, mother’s age at death^100^, childhood IQ^101^, college completion^96^, years of schooling^93^, age at menopause^102^, and age at first birth^97^. A likelihood ratio test implemented in R with the *anova* function was used to compare the goodness of fit of the joint MLR models with and without each individual PGS and p-values were used to determine significance of the variability in the mean PGS across groups (p_global_). Significance was declared after Bonferroni correction for 127 p-values (p<0.05/127=0.0004).

#### Extension cohort: The adolescent brain cognitive development (ABCD) Study

The Adolescent Brain and Cognitive Development (ABCD) study^48,103^ (http://abcdstudy.org) is an observational study following 11 875 children from across the US, starting at age nine or ten with limited exclusion criteria to create a socio-economically and demographically representative cohort. ABCD administers biennial assessments of physical health, mental health, neuro cognition, family, cultural and environmental variables, SUD, genetic and other biomarkers, and multi-modal neuroimaging. Genotyping used the Smokescreen array, imputations used the Michigan Imputation Server, and quality control is described in Loughnan et al.^104^. We used measures of 51 assessments (**Supplementary table 19**) covering categorical and dimensional psychopathology, mania symptoms, prodromal psychosis, impulsivity, and behavioral activation and inhibition collected at enrollment^48^. To protect against confounding by population structure, we focus on the homogenous, European ancestry sub-cohort of 5 455 children selected following genetic ancestry estimation using fastStructure^105^.

#### Polygenic Scores in ABCD

PGS in ABCD were calculated with PRSice^106^ and summary statistics from each of the three ADHD case variable GWAS described above. SNPs were pruned and clumped (clumping r^2^□=□0.1, distance□=□250□kb), but no threshold on p-value was set (i.e., all independent SNPs, p<1, were included). SNPs within the major histocompatibility complex were removed. Generalized linear models (GLMs) were used to test the association of each of the three PGS across each of 51 assessments, separately. GLMs included fixed effects of ADHD case-control PGS, age, sex at birth, data collection site, and 10 ancestry principal components. To control for family effects (twins and siblings within the sample), we iteratively fit 100 models for each behavior taking a random selection on singletons (down sampling to one individual from each family, n=4 622) and report the median across iterations. The distribution of each of the baseline assessment (response variable) was analyzed to ensure normality or to handle zero inflation and right skewness. Non-zero inflated distributions were rank normalized (as was performed for PGS) and the GLM was fitted using the family=gaussian option. For the right-skewed, zero-inflated distributions, data were shifted to ensure non-negativity and GLM were fitted with a gamma distribution and a log link function. Summary measures making up the Kiddie Schedule Disorders and for Schizophrenia (KSADS), except for the two KSADS Total Symptoms measures, were binarized using a median split and fit using logistic regression. This decision to binarize was due to model convergence issues resulting from very heavy distribution skews when attempting to fit using a gamma distribution. Bonferroni adjustment was used to declare significance (p<0.05/51=0.001) and nominally significant tests were noted suggestive (p<0.05).

## Data Availability

Data is available on request and in accordance with Danish law.

## Acknowledgments

Data used in the preparation of this article were obtained from the Adolescent Brain Cognitive Development (ABCD) Study (https://abcdstudy.org), held in the NIMH Data Archive (NDA). This is a multisite, longitudinal study designed to recruit more than 10,000 children age 9-10 and follow them over 10 years into early adulthood. The ABCD Study is supported by the National Institutes of Health and additional federal partners under award numbers U01DA041022, U01DA041028, U01DA041048, U01DA041089, U01DA041106, U01DA041117, U01DA041120, U01DA041134, U01DA041148, U01DA041156, U01DA041174, U24DA041123, and U24DA041147. A full list of supporters is available at https://abcdstudy.org/nih-collaborators. A listing of participating sites and a complete listing of the study investigators can be found at https://abcdstudy.org/principal-investigators.html. ABCD consortium investigators designed and implemented the study and/or provided data but did not necessarily participate in analysis or writing of this report. This manuscript reflects the views of the authors and may not reflect the opinions or views of the NIH or ABCD consortium investigators. The ABCD data repository grows and changes over time. The iPSYCH Initiative is funded by the Lundbeck Foundation (grant numbers R102-A9118 and R155-2014-1724), the Mental Health Services Capital Region of Denmark, University of Copenhagen, Aarhus University and the university hospital in Aarhus. Genotyping of iPSYCH samples was supported by grants from the Lundbeck Foundation, the Stanley Foundation, the Simons Foundation (SFARI 311789), and NIMH (5U01MH094432-02). The IPSYCH Initiative utilize the Danish National Biobank resource that is supported by the Novo Nordisk Foundation. IPSYCH data was stored and analyzed at the Computerome HPC facility (http://www.computerome.dtu.dk/) and authors are grateful for continuous support from the HPC team led by A. Syed of DTU Bioinformatics, Technical University of Denmark. AJS acknowledges support from Lundbeckfonden under the Fellowship R335-2019-2318 and the National Institute for Aging of the National Institutes of Health under awards U19AG023122, U24AG051129S1, UH2AG064706, and UH2AG064706S1. SLB acknowledges support from the Research Fund of the Mental Health Services – Capital Region of Denmark R4A92, The Lundbeck Foundation R208-2015-3951 and Fonden for Faglig Udvikling af Speciallægepraksis 38850/16. SD acknowledges research support from the European Commission (Horizon 2020, grant no 667302), Helsefonden (grant no 19-8-0260) and the European Union’s Horizon 2020 research and innovation programme under grant agreement No 847879.

## Author Contributions

The aims of this paper were conceived of jointly by S.L., I.B., S.D., T.M.W., and A.J.S and supervised broadly by S.D., T.M.W., A.J.S. The overall study design was developed by S.L., I.B., and A.J.S, with various components of the paper conducted with input and guidance from collaborators. Variable extraction from and definition in registers was conducted by S.L., I.B., and D.H., with assistance and guidance from E.A., M.G.P, and S.D. SNP heritability, genetic correlations, GWAS, and PGS generation were conducted by S.L. and A.J.S, with assistance from J.G, V.A., M.V., A.I., supervised by A.J.S. Functional annotations were conducted by S.L., with assistance, design, and supervision from R.W., D.G., and A.J.S. Single locus and polygenic multinomial tests were conducted by S.L., J.M., and A.J.S., using a statistical implementation from A.D. and N.Z with supervision from A.D., N.Z., and A.J.S. Analysis of ABCD was designed and conducted by R.L. and C.P., supervised by T.J. Simulations were conducted by A.J.S., with support from S.L., M.K., and K.K. A.D.B., D.M.H., O.M., M.N., P.B.M, T.M.W. contributed iPSYCH data. T.J. contributed ABCD data. The initial draft was written by S.L., with subsequent versions written by S.L., I.B., and A.J.S. All authors discussed results, commented on drafts, and provided critical feedback throughout.

