## Supplementary Note for "Polygenic profiles define aspects of clinical heterogeneity in ADHD"

**Supplementary Note:** On generative models and polygenic score profiles

In this manuscript we pursue a novel multinomial model for multivariate profiles of polygenic scores (PGS) to suggest a powerful, quantitative test of genetic heterogeneity in ADHD (Main text, Figure 4). The fundamental concept we explore is estimated and plotted in Figure 4 – that the mean *genetic values* (i.e., true, underlying genetic liability) for a number of traits may be different in selected subsets of individuals diagnosed with ADHD. Our statistical test includes extra layers of complexity, estimating differences across multiple groups simultaneously to include controls, in multiple scores jointly to handle collinearity and genetic correlation, and with covariates to protect against potential confounding due to ancestry differences, that are set aside for this note. Instead, we follow inspiration from Klein and Riso 1994^1^, who considered various generative models that may result in comorbidity of multifactorial disorders, and Neale and Kendler 1995^2^, who extended this to consider familial resemblance in psychiatric outcomes. We use simulations from a multivariate liability threshold model to illustrate expected 3-score PGS profiles for control vs. child ADHD vs. adult ADHD and for individual with or without each of ASD and ADHD, under various scenarios. These simulations are not intended to be exhaustive or definitive, but rather to demonstrate a few important points regarding the power and uniqueness of our approach and add extended discussion points for two important and timely findings relating to ADHD etiology. In particular, we hope to emphasize the following points with these simulations:

1) Our results are unlikely to be driven by simple misdiagnosis, simple models of genetic intensity related to age of onset, or simple accumulation of exposures.

2) Incorporating multiple PGS into our approach (i.e., being a multivariate model) is powerful and allows for more nuanced inferences than when considering primary disorder PGS (i.e., ADHD) alone.

3) Incorporating undiagnosed controls simultaneously with various diagnosed groups in our approach (i.e., being a multinomial model) gives an important, fuller context for case group differences.

4) Simple generative models based on infinitesimal, exchangeable models of pleiotropy (i.e., all SNPs are drawn from one distribution) may not be enough to explain observed pictures of genetic heterogeneity, and less parsimonious models should be considered (e.g., pathway models, local genetic correlations, etc.).

*Adult diagnosis of ADHD*

There are multiple potential explanations for an adult diagnosis of ADHD. It has only recently been incorporated into clinical guidelines and there are numerous debates as to its etiology. We consider five potential generative models, each of which being a variant of a multivariate liability threshold model, with results presented in supplementary figures 11 to 15.

For each model, we generate a truth population according to a multivariate liability model, where liability for each trait, *l_i_*, is normally distributed, and arises from the correlated, additive effects of genotypes, and an independent environmental component.

$$\boldsymbol{l}=\boldsymbol{\beta X}+\boldsymbol{\varepsilon}$$

$$\boldsymbol{X}\sim N(\boldsymbol{0,I})$$

$$\boldsymbol{\varepsilon\sim}N\left( \boldsymbol{0,}\left[ \begin{matrix} 1-h_{ADHD}^{2} & 0 & 0 \\ 0 & 1-h_{MDD}^{2} & 0 \\ 0 & 0 & 1-h_{EA}^{2} \end{matrix} \right] \right)$$

$$\boldsymbol{\beta}\sim N\left( \boldsymbol{0},\frac{1}{m}\left[ \begin{matrix} h_{ADHD}^{2} & \rho_{G, ADHD,MDD}\sqrt{h_{ADHD}^{2}h_{MDD}^{2}} & \rho_{G,ADHD,EA}\sqrt{h_{ADHD}^{2}h_{EA}^{2}} \\ \rho_{G,ADHD,MDD}\sqrt{h_{ADHD}^{2}h_{MDD}^{2}} & h_{MDD}^{2} & \rho_{G,MDD,EA}\sqrt{h_{MDD}^{2}h_{EA}^{2}} \\ {\rho_{G,ADHD,EA}}\sqrt{h_{ADHD}^{2}h_{EA}^{2}} & \rho_{G,MDD,EA}\sqrt{h_{MDD}^{2}h_{EA}^{2}} & h_{EA}^{2} \end{matrix} \right] \right)$$

Diagnoses are modeled according to a thresholding of this underlying liability. Diagnoses are assigned when the liability for trait, *t*, in individual, *i*, is above a threshold defined by the inverse of the normal cumulative distribution function (CDF) chosen such that cases reflect the upper tail proportion, *k_t_*.

$${dx}_{t,i}=I\left( l_{t,i}>\Phi^{-1}\left( 1-k_{t} \right) \right)$$

For simulated ADHD and MDD we set the prevalence, *k_t_*, as 0.05 and 0.15, respectively. For ADHD, MDD, and educational attainment (EA) we defined the narrow sense heritability (*h^2^*) to be 0.7, 0.45, and 0.4, and the SNP-heritability (*h^2^_SNP_*) to be 0.2 for each. We defined the genetic correlations (𝜌_G_) between ADHD and MDD, ADHD and EA, and MDD and EA to be 0.4, -0.5, and -0.25, respectively. For all simulations, a population size of 100,000 was used, and the number of independent genotypes, *m*, was set to 250**.** Multivariate, correlated genetic effects,$\boldsymbol{\beta}$, were generated by transforming independent, normal variables, B~N(0,I), according to ***BU^T^***, where **U^T^** is the Cholesky decomposition of the genetic correlation matrix, ***P_G_=U^T^U***, obtained using the R function chol(). Genetic effects were scaled to satisfy the expected heritability. Simulations were performed twice for each scenario, once using the narrow sense heritability, and one using the SNP-heritability.

*Model 0: Null*. As a null model we consider the age of diagnosis to be a random feature of ADHD.

For each individual, we drew an age of onset from a uniform distribution over the set *a*={5,10,15,20}. We calculated the means of the true genetic values, ***βX***, for each trait, in the three groups of interest: child onset ADHD (*dx_ADHD_*=1 and *a* in {5,10,15}), and for adult onset ADHD, (*dx_ADHD_*=1 and *a* in {20}), and individuals without ADHD, (*dx_ADHD_*=0).

The results are shown in Supplementary Figure 11. The illustrated genetic value distributions are the same for adult and child diagnosis, and show the same differences from controls, as expected, but inconsistent with our results in main text Figure 4.

*Model 1: Genetic Severity*. Often age of onset is considered to be a sign of genetic severity and thus it has been suggested that early age of diagnosis may be related to increased genetic burden.

To model this, for each individual, we drew an age of diagnosis, *a*, from a different set of values depending on their genetic liability for ADHD. ADHD cases in the lowest quartile of genetic liability had *a* drawn with uniform probability from the set {20,20,20,15,10,5}, in the second quartile from the set {20,15,15,15,10,5}, in the third quartile from the set {20,15,10,10,10,5}, and in the highest quartile from the set {20,15,10,5,5,5}. This created a negative association between ADHD liability and age of diagnosis (i.e, more ADHD burden, lower age of diagnosis).

The results are shown in Supplementary Figure 12. The illustrated genetic value distributions are different for adult and child diagnosis, but also inconsistent with our results in main text Figure 4. Interestingly, the expected trend regarding ADHD PGS is consistent with our results (green arrow), but discrepancies appear when considering secondary PGS of MDD and EA which show the opposite trend (red arrows). This is a function of the population genetic covariance and demonstrates the utility of considering multiple PGS profiles.

*Model 2: Persistence*. Because diagnostic manuals require a retrospective report of ADHD symptoms during childhood for an adult diagnosis, and because many individuals see their symptoms remit with age, an alternate argument is that, unlike with other disorders, genetically more severe cases might be marked by persistence into adulthood. Under this view, a first diagnosis as an adult would be capturing persistent cases missed during childhood.

Here, for each individual, we drew an age of onset from a different set of values depending on their genetic liability for ADHD, in the opposite way from above. ADHD cases in the *highest* quartile of genetic liability had *a* drawn with uniform probability from the set {20,20,20,15,10,5}, in the *third* quartile from the set {20,15,15,15,10,5}, in the *second* quartile from the set {20,15,10,10,10,5}, and in the *lowest* quartile from the set {20,15,10,5,5,5}.

The results are shown in Supplementary Figure 13. The illustrated genetic values are also inconsistent with our results in main text Figure 4. Here, the expected trend regarding ADHD PGS is inconsistent with our results (adult ADHD higher than child ADHD, red arrow), while the trends with respect secondary PGS of MDD and EA appear more qualitatively consistent (green arrows).

*Model 3: Misdiagnosis*. The nature of adult ADHD, while incorporated into clinical diagnostic manuals, is still debated among researchers. The idea that adult ADHD could be truly marked by symptom onset in adulthood is an especially strong point of controversy. One explanation for a diagnosis without persistence from childhood could be, then, that the symptoms of another disorder are mistaken for ADHD.

Here, we consider our null hypothesis of age of diagnosis being random with respect to ADHD liability. Motivated by the increased PGS for MDD and depressive symptoms observed in adult diagnosed ADHD, we then assigned a random sample of individuals above the MDD threshold an ADHD diagnosis. Here, our adult ADHD sample was 80% individuals selected as above the ADHD liability threshold and 20% individuals selected as above the MDD threshold.

The results are shown in Supplementary Figure 14. Interestingly, these results illustrate trends that are similar in terms of ADHD and MDD values (green arrows), but different in terms of EA value (red arrow). The genetic correlation between ADHD and EA is more strongly negative than between MDD and EA, and so adding misdiagnosed MDD patients makes the mean EA value of the adult ADHD group less extreme, again demonstrating how, given a population genetic covariance, we can leverage multiple scores for inference.

*Model 4: Education as an exposure.* Model 1, where childhood diagnosis is more genetically severe, is conceptually equivalent to models where random environmental effects accumulate over a lifespan to compensate for an individual’s (lack of) genetic risk factors. Here, we consider a slightly different model, whereby as an individual ages the negative of their EA value contributes to their ADHD liability. The idea here is that an individual who has not onset ADHD may be more or less likely to develop ADHD based on their educational attainment, with low attainment becoming activated as an age dependent risk factor.

For this model, we assigned a random age of *potential* diagnosis to each individual (i.e., a uniform draw from the set {5,10,15,20}, whether or not they were above the ADHD liability threshold). For individuals assigned a potential age of diagnosis as 20, we added the negative of their EA liability to their ADHD liability, and then called the diagnoses according the updated liability and previously defined threshold.

These results are presented in Supplementary Figure 15. As with model 3, these are consistent in two of the three trends (green arrows), ADHD and EA values, but different in terms of MDD values (red arrow). Here, because the negative population genetic correlation between EA and MDD is modest, and more negative than the EA-ADHD correlation, adding the EA component to the ADHD liability reduces the expected MDD value in those diagnosed with ADHD.

*Conclusions.* Regarding an adult diagnosis of ADHD, we hope to have emphasized a few important points. First, our results presented in Figure 4 are not consistent with simply an MDD misdiagnosis or increasing environmental exposure. Second, and importantly, these inferences could not be made without a broader consideration of adjacent (i.e., non-ADHD) genetic values. Models 1, 3, and 4 described here would all be consistent with reduced ADHD PGS in those diagnosed as an adult, but, given the proposed population genetic covariance, considering even simple profiles of scores can potentially distinguish among confounding or simple models. Thirdly, more flexible, less parsimonious models than posed here are needed to generate the differences in profiles. These models could include a genetic heterogeneity arising from SNP group or pathway specific genetic correlation, mixtures of factors, a reconsideration of our population parameters, or the potential of systematic bias or error in PGS. Exploring all of those are beyond the scope of our work here, which focuses on an approach for detecting heterogeneity using PGS profiles rather than reconsidering observed population genetic parameters.

*ADHD adjacent ASD*

Here we follow the same procedure as above for generating illustrative genetic value profiles, but consider profiles for ADHD, ASD, and EA, and describe the mean genetic values for individuals diagnosed with both, either, or neither disorder under different population models in supplementary figures 16 to 24.

For our simulated ADHD and ASD we set the prevalence, *k*, as 0.05 and 0.01, respectively. *h^2^* for ADHD, ASD, and EA we chosen as 0.7, 0.8, and 0.4, with *h^2^_SNP_* as 0.2 for each. Genetic correlations (𝜌_G_) between ADHD and EA, and ASD and EA to be -0.5, and 0.2, respectively. When modeling ASD and ADHD liabilities according to a correlated liability model, 𝜌_G_ was set to 0.33, and 0 otherwise, keeping the other trait 𝜌_G_ the same.

*Model 0: Null.* As a null model we consider individuals with two diagnoses to be those who happen to be above the liability threshold for both disorders, simultaneously. Here, we did not model the liabilities for ADHD and ASD as correlated. The results of this simulation are shown in Supplementary Figure 16 and actually recapitulate trends depicted in Figure 4 rather well, with the double diagnosed cases appearing similar to each single diagnosed case in terms of ADHD and ASD PGS, despite fixing the ADHD-ASD 𝜌_G_ to 0 in the population.

*Model 1: Correlated liability.* Here we generate diagnostic status as in Model 0, but under the assumption that ADHD and ASD liability are genetically correlated. Surprisingly, the results (Supplementary Figure 17) here look more discrepant, as the single diagnosed cases may be expected to have an increase value over controls, and the double diagnosed cases increased values over the single diagnosed cases. The strongest point of departure is apparent when considering controls relative to the various case-groups, which is an important strength of our multinomial approach.

*Model 2. ASD misdiagnosis.* An excess of comorbidity over that expected from bivariate liability can arise because of misdiagnosis. Misdiagnosis can also inflate genetic correlations in the population. Here consider one-way misdiagnosis and we assign a random 10% of ADHD cases, an ASD diagnosis. This is expected to result in an observed genetic correlation of ~0.3 under an uncorrelated liability model of the population (simulations not shown). These results, presented in Supplementary Figure 18, show, under these population parameters, genetic values among groups that are inconsistent with our results. The misdiagnosis reduced the mean ASD genetic value in doubly diagnosed cases substantially.

*Model 3. ADHD misdiagnosis.* Here we assign a random 80% of ASD cases, an ADHD diagnosis, which is expected to result in an observed genetic correlation of ~0.2 under an uncorrelated liability model of the population (simulations not shown). This extreme misdiagnosis rate is necessary to drive the observed genetic correlation towards a value observed in real data because the prevalence of the disorders is different. This makes this scenario significantly less plausible to begin with and the results in Supplementary Figure 19 are likewise inconsistent with our main text results, as the doubly diagnosed ADHD cases have their mean ADHD values driven significantly down towards the population mean.

*Model 4. Two-way misdiagnosis.* Here we assign a random 10% of ASD cases, an ADHD diagnosis, and vice versa, which is expected to result in an observed genetic correlation of ~0.3 under an uncorrelated liability model of the population (simulations not shown). Supplementary Figure 20, due to the prevalence of ADHD and ASD in the generated population, show very similar results to model 2.

*Model 5. Two-way misdiagnosis, correlated liability.* Here we assign a random 10% of ASD cases, an ADHD diagnosis, and vice versa, but from a population where ASD and ADHD are generated from *correlated* liabilities. This substantially inflated the observed genetic correlation from the generative 0.33 to ~0.6 (simulations not shown). Supplementary Figure 21 shows these results. The prevalence differences in the disorders means the mean ASD value among the double diagnosed cases is again driven down to a level beyond what we have observed.

*Model 6. Two-way causation.* This is a similar model to two-way misdiagnosis, except that the second diagnosis is a function of the second trait liability. Here we lower the threshold for a diagnosis of ADHD, if the individual is above the threshold for ASD, and vice versa. We increase the prevalence of each disorder, in the context of the other, buy a factor of 5, such that the ASD threshold for individuals with ADHD is defined by a different prevalence, 5*0.01=0.05, and for ADHD within ASD by 5*0.05=0.25. Supplementary Figure 22 shows these results for the uncorrelated liability baseline population. Lower the threshold in this way means that single diagnosed ADHD individuals end up having lower ASD value than the undiagnosed population, and single diagnosed ASD ends up with lower ADHD values. These trends are inconsistent with our results in the main text Figure 4.

*Model 7. Two-way causation, correlated liability.* We repeat model six in the population where ASD and ADHD liability have a 𝜌_G_ of 0.33. Here the results in Supplementary Figure 23 comport somewhat well with the observed trends in main text Figure 4.

*Model 8. Biological hierarchy.* Prior to the introduction of DSM-V, ADHD and ASD were not to be diagnosed simultaneously, with an ASD diagnosis being exclusionary for ADHD. This leads to a somewhat odd scenario in our simplistic simulations, where individuals above the threshold for ASD, should not have ADHD, regardless of their liability value. To model this scenario, we remove all ADHD diagnosed in the context of ASD. Because dual diagnosis exists in the real world, we reassign ADHD diagnosis to a random proportion of the ASD diagnosed individuals. Supplementary figure 24 depicts the genetic values from this somewhat contrived scenario, which do not fit observed trends well, as double diagnosed individuals resemble single diagnosed ASD cases, by construction.

*Conclusions.* The intersection of observed comorbidity, published estimates of modest genetic correlation, changing clinical practice, and the possibility of misdiagnosis between ADHD and ASD creates a scenario with many free parameters than can compensate for expected trends in genetic values by each parameter. Many of the differences illustrated in this set of simulations are more quantitative, as opposed to qualitative changes in trend directions above, which can challenge a clear interpretation. We still find importance in a few trends, however. For example, the models where comorbidity emerges due to a decoupling of genetic values in trait liability from that trait’s diagnosis (e.g., 2,3,4,5,6, and 8) describe the observed trends less well. This is important because it offers support for true comorbidity of disorder states among this dual diagnosed group. It is harder to distinguish between an uncorrelated liability model (i.e., model 0) and a correlated liability model with two-way causation (i.e., model 7) mutually lowering thresholds for a diagnosis on the basis of average genetic values, although the former is inconsistent with reports of correlated genetic liability between ADHD and ASD. Here, unlike the previous scenario, our multivariate profile approach aids in inference (low support for models 2, 5, or 6) by including controls, demonstrating strength of the multinomial model in our approach. Again, we are limited here by our choice to work with population parameters that more or less reflect the current state-of-the-field opinions. Some of the scenarios we propose (e.g., misdiagnosis) could permeate the studies that have painted this state-of-the-field and could contribute to simulation-result differences or represent alternative scenarios we have not considered.

1	Klein, D. N. & Riso, L. P. in *Basic issues in psychopathology*	(ed C. G. Costello) 19-66 (Guilford Press, 1993).

2	Neale, M. C. & Kendler, K. S. Models of comorbidity for multifactorial disorders. *Am J Hum Genet* **57**, 935-953 (1995).
