## Supplementary Figures for "Polygenic profiles define aspects of clinical heterogeneity in ADHD"

**Supplementary figure 1.** Hierarchical clustering of tetrachoric correlations computed for 22 ADHD-adjacent traits

**Supplementary figure 2.** Manhattan and QQ plots for GWAS of First ADHD dx as an adult

**Supplementary figure 3.** Manhattan and QQ plots for GWAS of ADHD-adjacent SUD

**Supplementary figure 4.** Manhattan and QQ plots for GWAS of ADHD-adjacent ASD

**Supplementary figure 5.** PheWAS for rs8178395 and rs8178289 using GWAS Atlas

**Supplementary figure 6.** LD comparisons with 1KGP

**Supplementary figure 7.** MAF comparisons with 1KGP

**Supplementary figure 8.** LDSC regression partitioned heritability for First ADHD dx as an adult

**Supplementary figure 9.** LDSC regression partitioned heritability for ADHD-adjacent SUD

**Supplementary figure 10.** LDSC regression partitioned heritability for ADHD-adjacent ASD

**Supplementary figure 11.** Adult diagnosed ADHD, simulations, model 0: Null.

**Supplementary figure 12.** Adult diagnosed ADHD, simulations, model 1: Genetic severity.

**Supplementary figure 13.** Adult diagnosed ADHD, simulations, model 2: Persistence.

**Supplementary figure 14.** Adult diagnosed ADHD, simulations, model 3: Misdiagnosis.

**Supplementary figure 15.** Adult diagnosed ADHD, simulations, model 4: Educational exposure.

**Supplementary figure 16.** ADHD adjacent ASD, simulations, model 0: Null.

**Supplementary figure 17.** ADHD adjacent ASD, simulations, model 1: Correlated liability.

**Supplementary figure 18.** ADHD adjacent ASD, simulations, model 2: ASD misdiagnosis.

**Supplementary figure 19.** ADHD adjacent ASD, simulations, model 3: ADHD misdiagnosis.

**Supplementary figure 20.** ADHD adjacent ASD, simulations, model 4: Two-way misdiagnosis.

**Supplementary figure 21.** ADHD adjacent ASD, simulations, model 5: Two-way misdiagnosis, correlated liability.

**Supplementary figure 22.** ADHD adjacent ASD, simulations, model 6: Two-way causation.

**Supplementary figure 23.** ADHD adjacent ASD, simulations, model 7: Two-way causation, correlated liability

**Supplementary figure 24.** ADHD adjacent ASD, simulations, model 8: Biological hierarchy.

**Supplementary figure 25.** Single PGS multinomial logistic regression analysis for the three ADHD-adjacent traits displaying PGS levels

**Supplementary figure 26.** Single PGS multinomial logistic regression analysis for the three ADHD-adjacent traits displaying contrast effect estimates

**Supplementary figure 27.** Joint PGS multinomial logistic regression analysis for the three ADHD-adjacent traits displaying contrast effect estimates

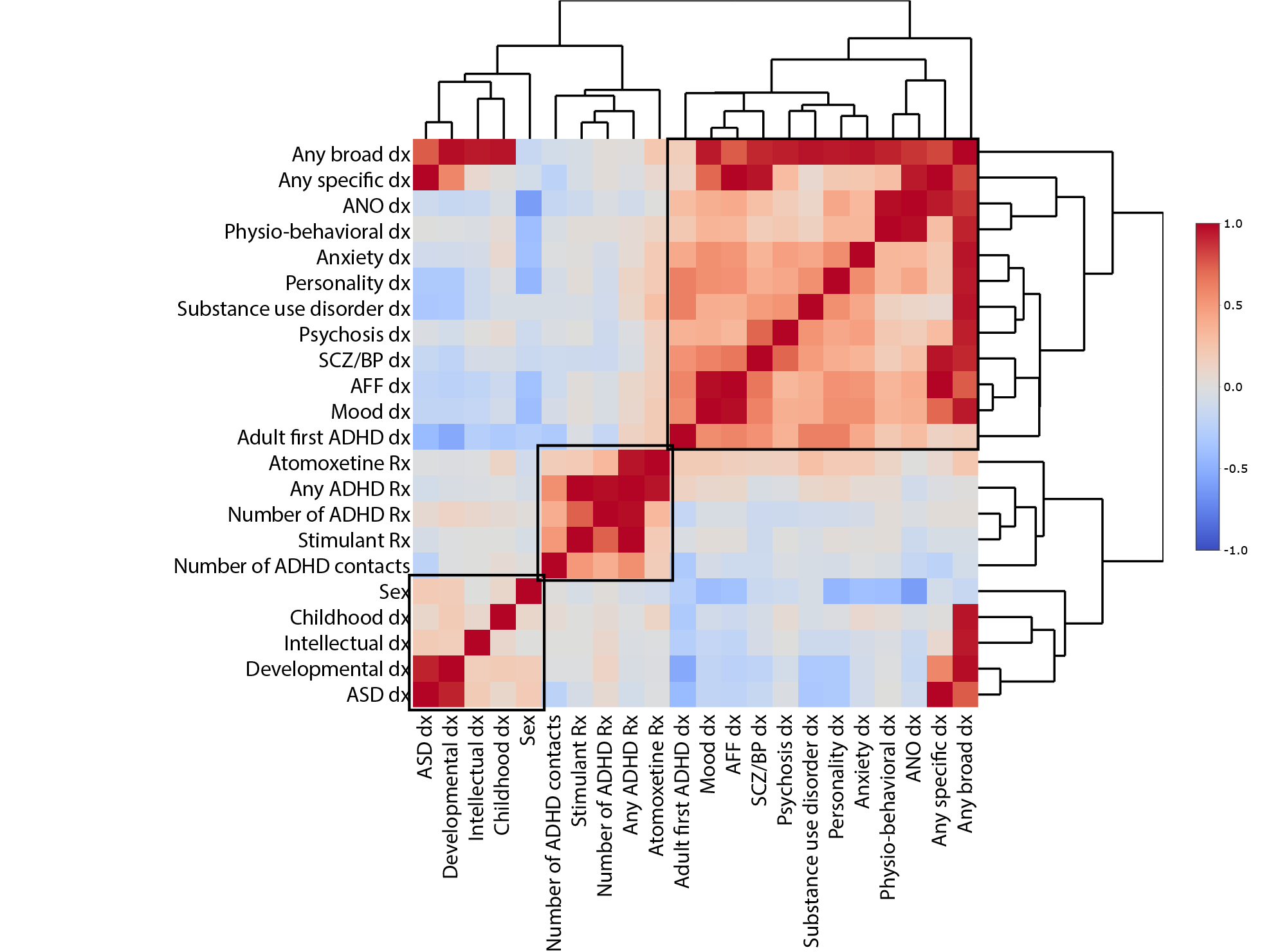

**b**

**Supplementary figure 1**. **Hierarchical clustering of pairwise tetrachoric correlations of ADHD-adjacent traits reveals some latent structure.** We clustered the 22 ADHD-adjacent traits on the basis of tetrachoric correlations using the *heatmaply* package in R. Roughly three clusters of ADHD-adjacent traits. Numeric values are reported for all correlation pairs in Supplementary table 3.

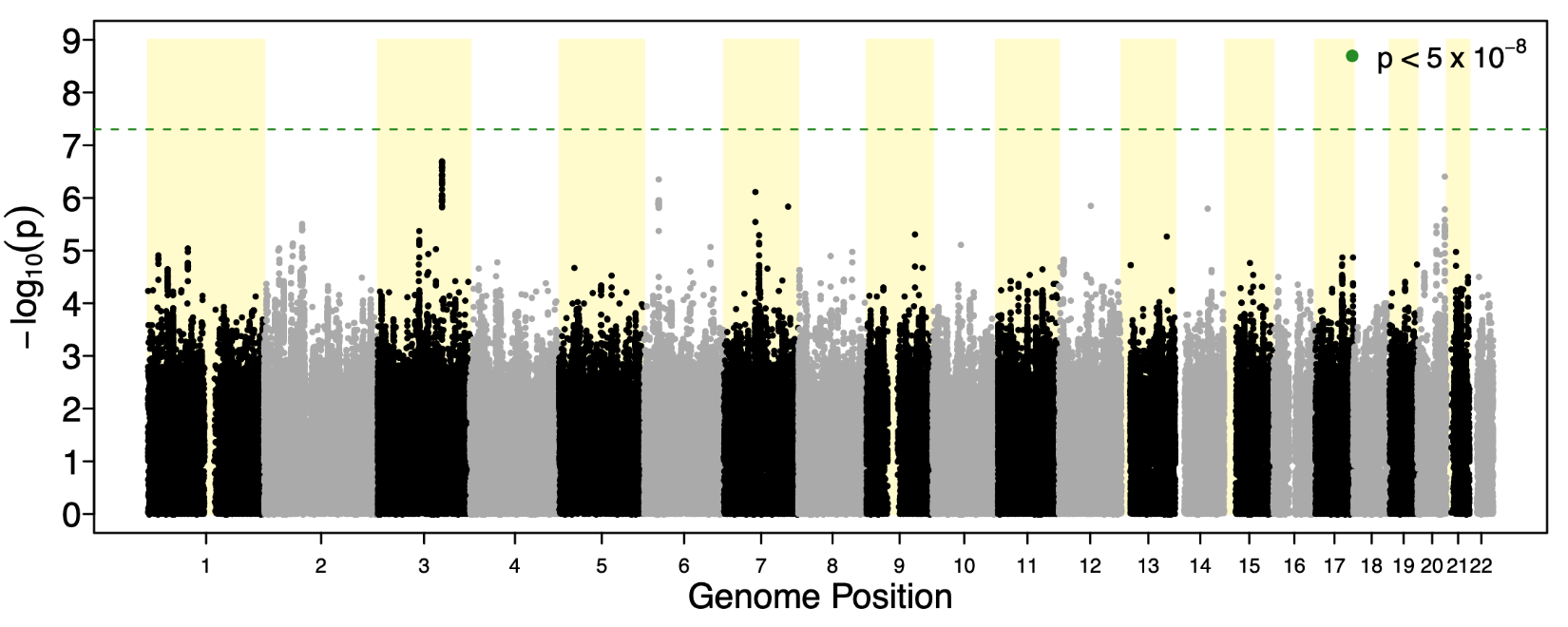

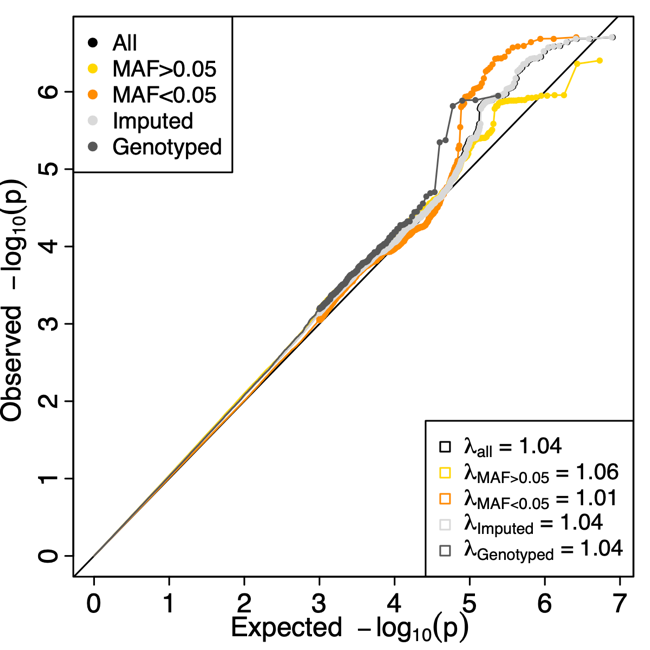

**Supplementary figure 2.** Manhattan and QQ-plot for first diagnosis of ADHD ≥18 years (First ADHD dx as an adult ). The left panel depicts the –log10 of two-sided p-values (y-axis) from logistic regression association tests performed in GWAS (n= 3 323 cases with adult ADHD Dx, n= 10 761 cases with childhood ADHD Dx) of 8 019 760 SNP dosages depicted at genome position (x-axis). The horizontal dotted green line represents the significance threshold at p<5x10^-8^. The right panel shows the quantile-quantile (QQ) plot comparing the distribution of –log10 association P values to that expected under the global null hypothesis. Depicting all SNPs (All), common (MAF>0.05), rare (MAF <0.05), imputed only (Imputed), genotyped only (Genotyped) and their estimated genomic inflation factor (λ)

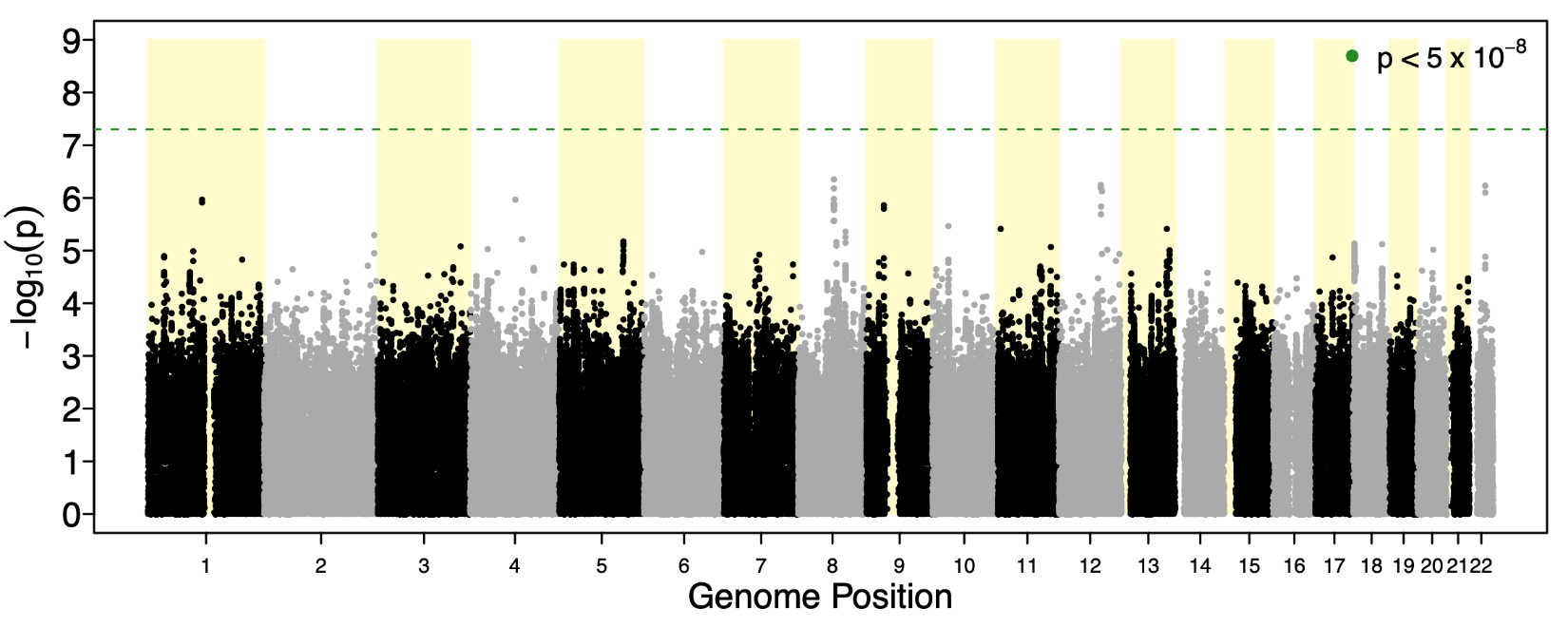

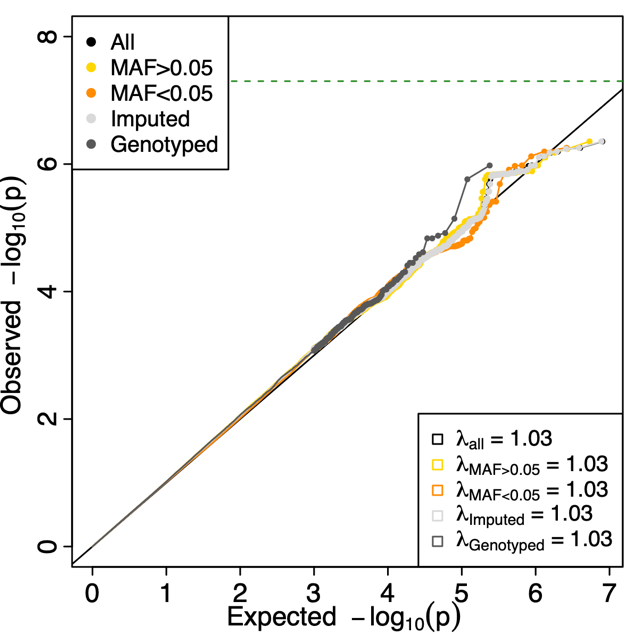

**Supplementary figure 3.** Manhattan and QQ-plot for ADHD-adjacent diagnosis of substance use disorder (SUD dx). The left panel depicts the –log10 of two-sided p-values (y-axis) from logistic regression association tests performed in GWAS (n=2 627 ADHD cases with substance use, n=11 457 ADHD cases without substance use) of 8 019 760 SNP dosages depicted at genome position (x-axis). The horizontal dotted green line represents the significance threshold at p<5x10^-8^. The right panel shows the quantile-quantile (QQ) plot comparing the distribution of –log10 association P values to that expected under the global null hypothesis. Depicting all SNPs (All), common (MAF>0.05), rare (MAF <0.05), imputed only (Imputed), genotyped only (Genotyped) and their estimated genomic inflation factor (λ)

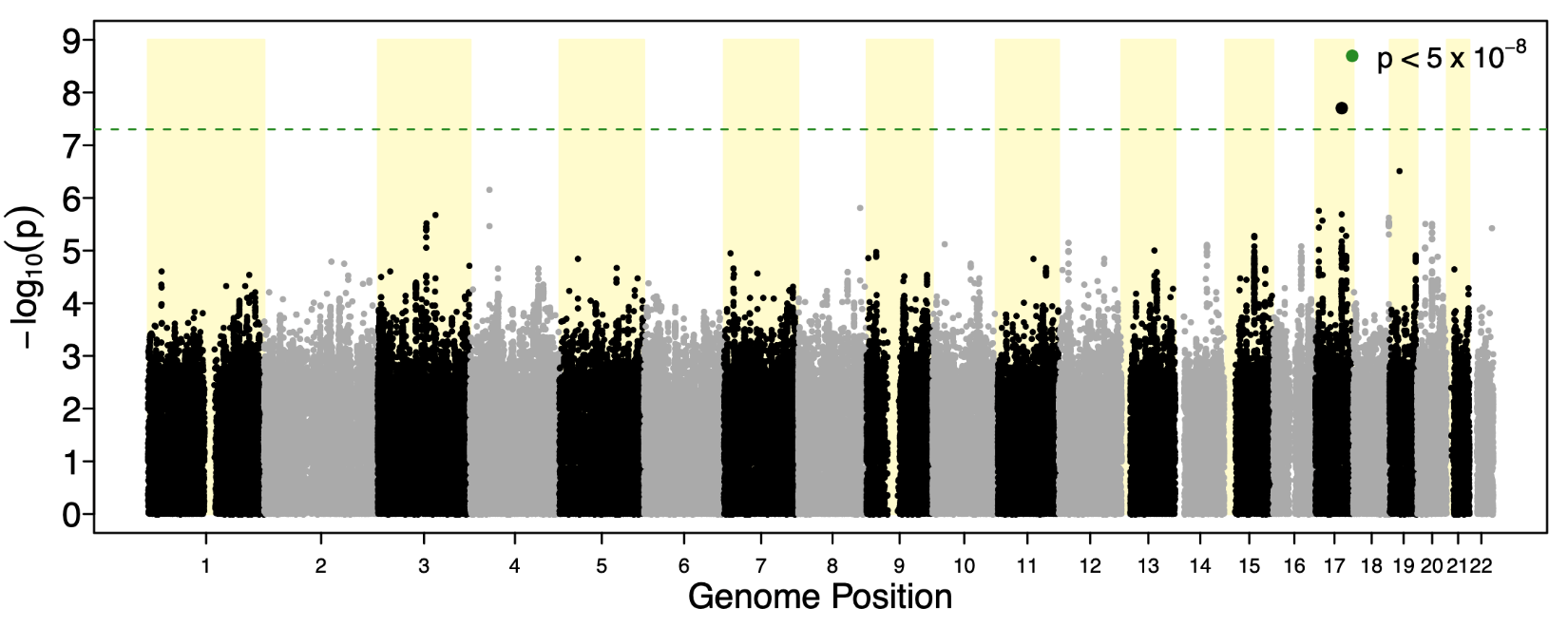

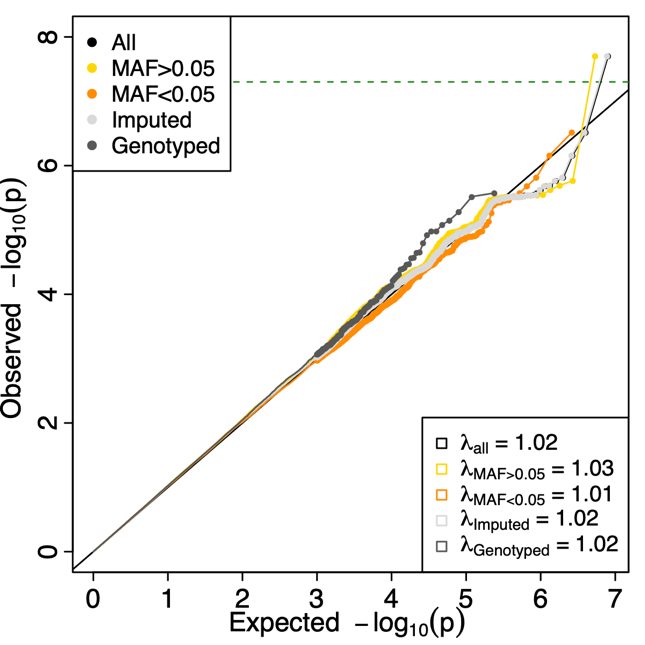

**Supplementary figure 4.** Manhattan and QQ-plot for ADHD–adjacent diagnosis of ASD (ASD dx). The left panel depicts the –log10 of two-sided p-values (y-axis) from logistic regression association tests performed in GWAS (n=2 284 ADHD cases with ASD, n=11 800 ADHD cases without ASD) of 8 019 760 SNP dosages depicted at genome position (x-axis). The horizontal dotted green line represents the significance threshold at p<5x10^-8^. One genome wide significant locus is identified. The right panel shows the quantile-quantile (QQ) plot comparing the distribution of –log10 association P values to that expected under the global null hypothesis. Depicting all SNPs (All), common (MAF>0.05), rare (MAF <0.05), imputed only (Imputed), genotyped only (Genotyped) and their estimated genomic inflation factor (λ)

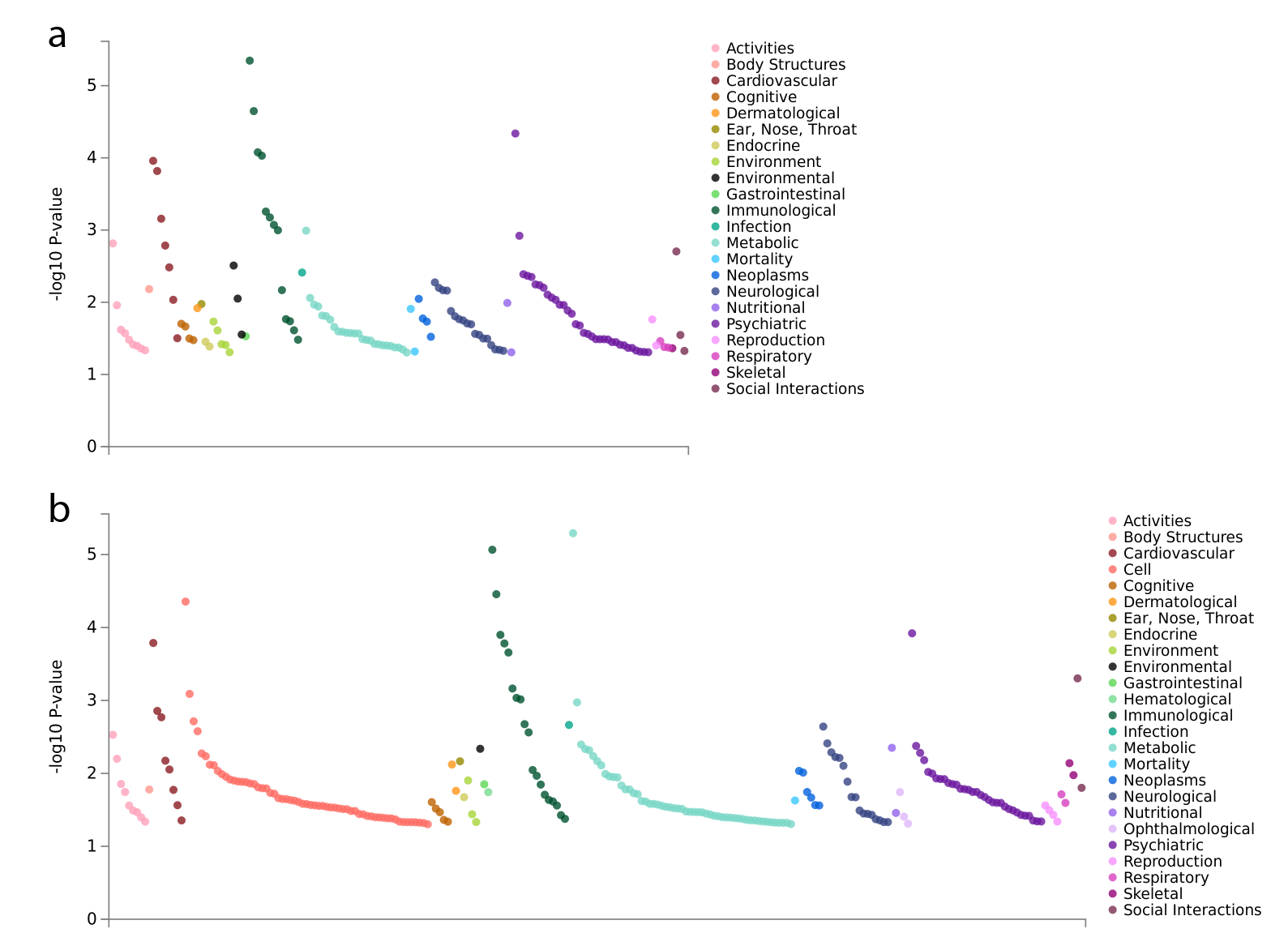

**Supplementary figure 5.** Phenome-wide association studies for **a**, lead SNP rs8178395 and **b**, proxy SNP rs8178289 performed using the GWASatlas web service (<https://atlas.ctglab.nl/PheWAS>). These show plausible prior associations with physiological measures of blood cell composition (Immunological), protein levels (Cell), and metabolites (Metabolic), cardiovascular complications (Cardiovascular), and sleep behavior (Psychiatric, see Supplementary table 5 for details).

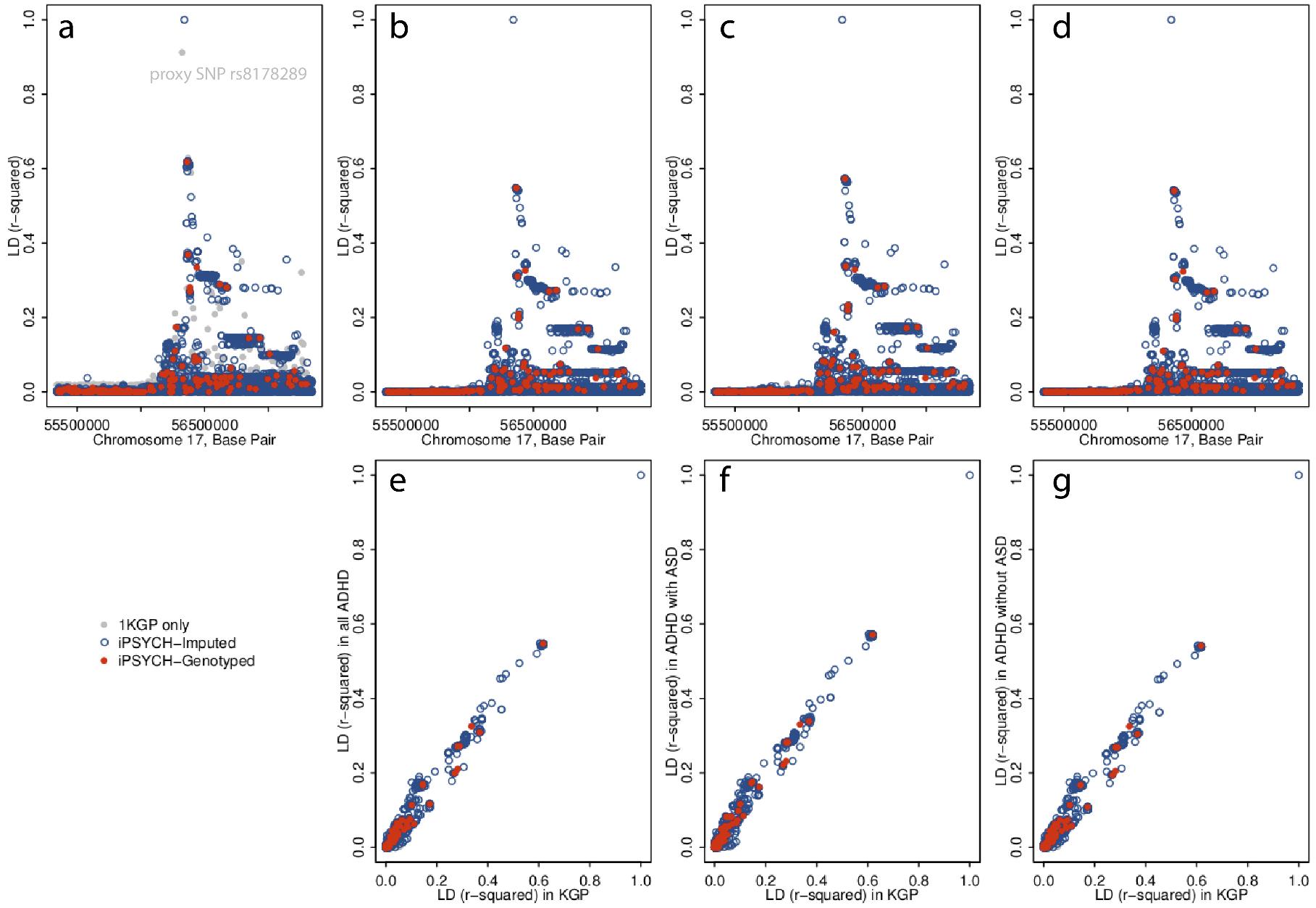

**Supplementary figure 6.** Comparison of local patterns of LD between rs8178395 and surrounding SNPs measured in the 1KGP and iPSYCH data. **a**, r-squared LD patterns in the locus surrounding rs8178395 as measured in the 1000 genomes project Europeans. **b**, The LD measures in all iPSYCH ADHD cases, **c**, in the iPSYCH ADHD cases with adjacent ASD, and **d**, in the iPSYCH ADHD cases without ASD are qualitatively similar, suggesting the sparse LD block is a function of this genomic region and not a potential genotyping or imputation error. The correlations between pairwise LD measured in the 1KGP and **e**, all iPSYCH ADHD cases, **f**, in the iPSYCH ADHD cases with adjacent ASD, and **g**, in the iPSYCH ADHD cases without ASD are strong with no outliers.

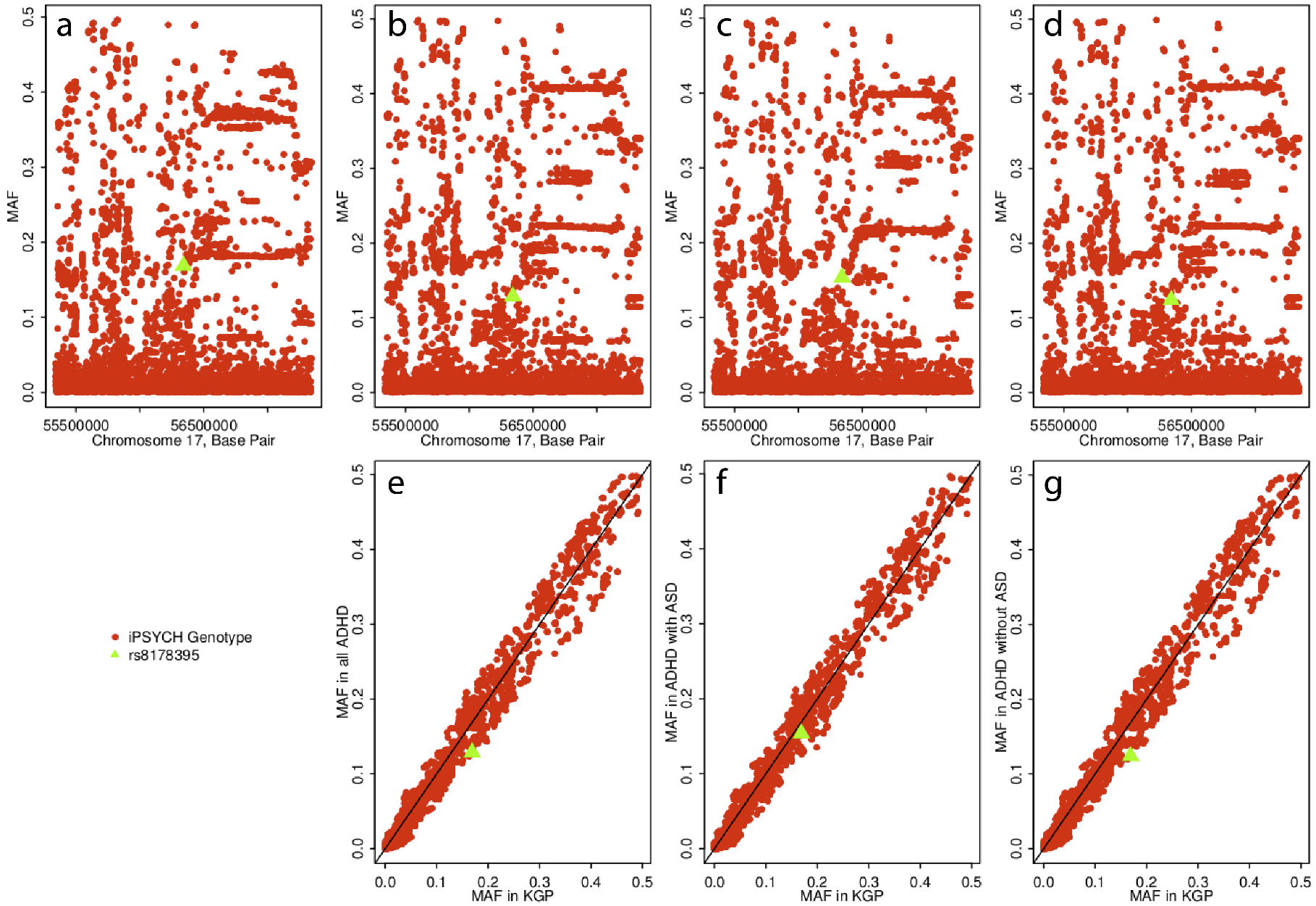

**Supplementary figure 7.** Comparison of MAF at rs8178395 and surrounding SNPs measured in the 1KGP and iPSYCH data. **a**, MAF patterns in the locus surrounding rs8178395 as measured in the 1000 genomes project Europeans. **b**, MAF measures in all iPSYCH ADHD cases, **c**, in the iPSYCH ADHD cases with ASD, and **d**, in the iPSYCH ADHD cases without ASD are qualitatively similar, suggesting a reasonably low potential of genotyping or imputation error. The correlations between MAF measured in the 1KGP and **e**, all iPSYCH ADHD cases, **f**, in the iPSYCH ADHD cases with ASD, and **g**, in the iPSYCH ADHD cases without ASD are strong with no outliers.

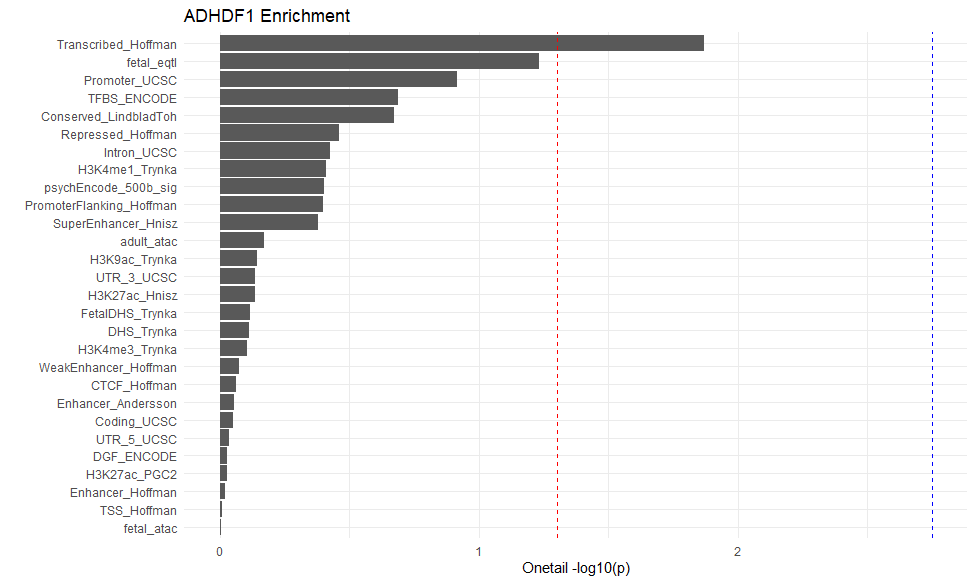

**Supplementary figure 8.** Enrichment categories for first diagnosis of ADHD as an adult (First ADHD dx as an adult ) ordered by significance: red line = -log10(0.05) & blue line = -log10(0.05/28) as estimated in partitioned LD score regression. The categories include fetal and adult brain eQTLs and ATAC, and a full baseline model as used in *Finucane et al. 2015*: Coding, 3' UTR, 5' UTR, promoter and intron: Digital genomic footprint and transcription factor binding site annotations: Combined chromHMM and Segway annotations for six cell: CTCF, promoter-flanking, transcribed, transcription start site (TSS), strong enhancer and weak enhancer categories are each a union over the six cell lines: DHSs: Cell type–specific H3K4me1, H3K4me3 and H3K9ac: Cell type–specific H3K27ac: Super-enhancers: Regions conserved in mammals: FANTOM5 enhancers. See Supplementary table 9 for numeric values of estimated enrichment.

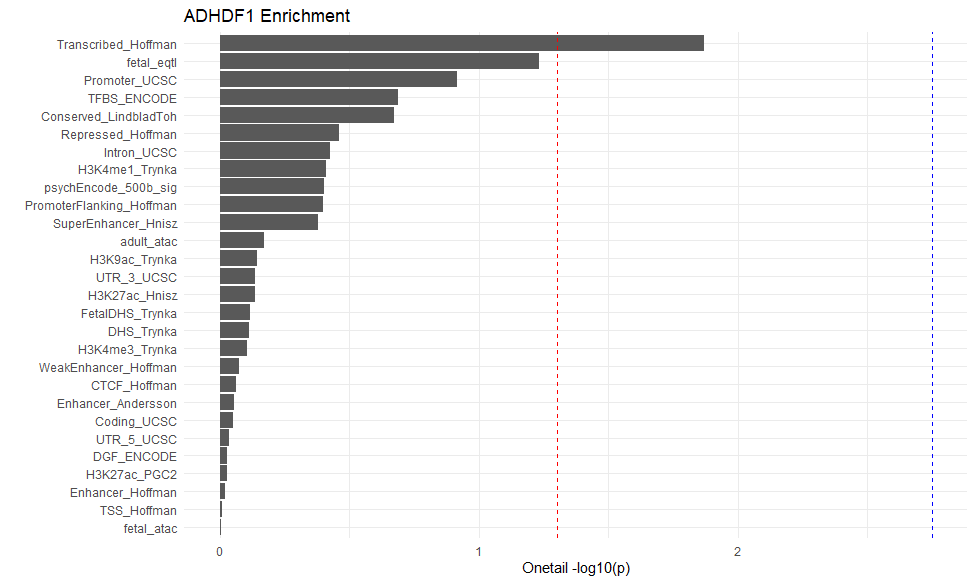

**Supplementary figure 9.** Enrichment categories for ADHD-adjacent diagnosis of substance use disorder (SUD dx) ordered by significance: red line = -log10(0.05) & blue line = -log10(0.05/28) as estimated in partitioned LD score regression. The categories include fetal and adult brain eQTLS and ATAC, a full baseline model as used in *Finucane et al*. *2015*: Coding, 3' UTR, 5' UTR, promoter and intron: Digital genomic footprint and transcription factor binding site annotations: Combined chromHMM and Segway annotations for six cell: CTCF, promoter-flanking, transcribed, transcription start site (TSS), strong enhancer and weak enhancer categories are each a union over the six cell lines: DHSs: Cell type–specific H3K4me1, H3K4me3 and H3K9ac: Cell type–specific H3K27ac: Super-enhancers: Regions conserved in mammals: FANTOM5 enhancers. See Supplementary table 9 for numeric values of estimated enrichment.

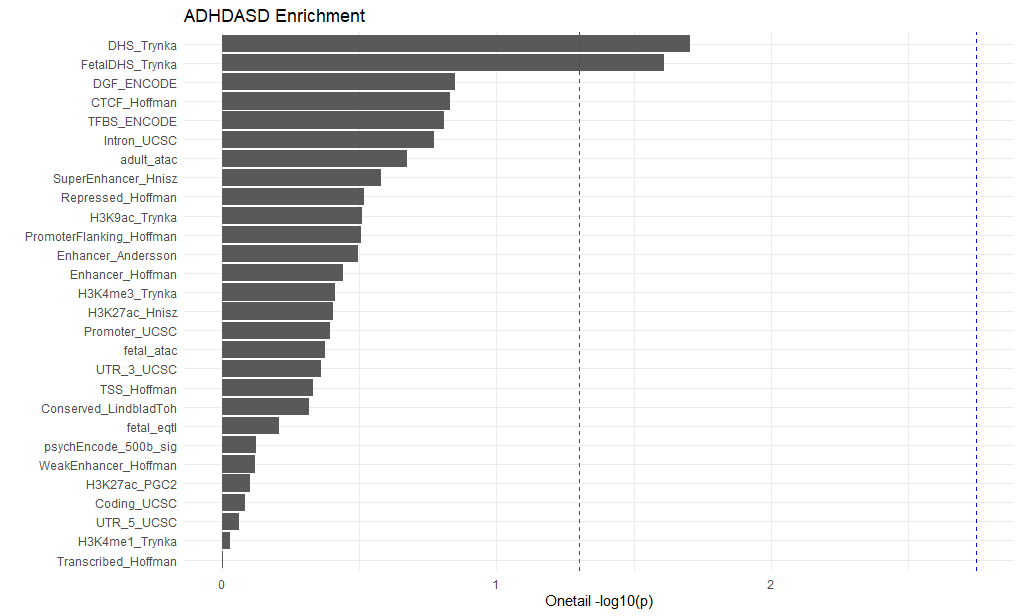

**Supplementary figure 10.** Enrichment categories for ADHD-adjacent diagnosis of ASD (ASD dx) ordered by significance: red line = -log10(0.05) & blue line = -log10 (0.05/28) as estimated in partitioned LD score regression. The categories include fetal and adult brain eQTLS and ATAC, a full baseline model as used in *Finucane et al. 2015*: Coding, 3' UTR, 5' UTR, promoter and intron: Digital genomic footprint and transcription factor binding site annotations: Combined chromHMM and Segway annotations for six cell: CTCF, promoter-flanking, transcribed, transcription start site (TSS), strong enhancer and weak enhancer categories are each a union over the six cell lines: DHSs: Cell type–specific H3K4me1, H3K4me3 and H3K9ac: Cell type–specific H3K27ac: Super-enhancers: Regions conserved in mammals: FANTOM5 enhancers. See Supplementary table 9 for numeric values of estimated enrichment.

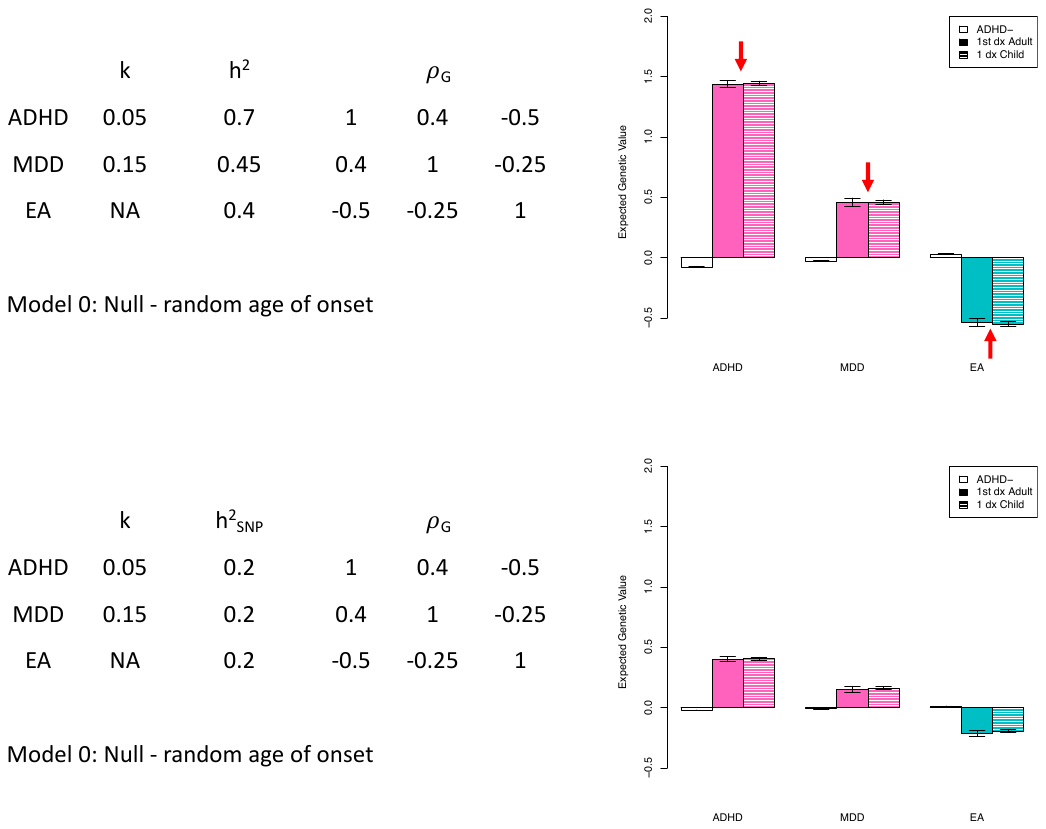

**Supplementary figure 11.** Adult diagnosis simulations model 0: random age of diagnosis. Here we use a multivariate, threshold liability model to generative illustrative genetic value profiles from a scenario that could underlie an adult versus child diagnosis of ADHD. These plots can be compared against main text Figure 4 to note qualitative differences (red arrows) or consistency (green arrows). On the left we show the population parameters that define the simulated population. Here, the age of diagnosis was chosen as a uniform probability draw from the set {5,10,15,20}. More methodological details and discussion are provided in the supplementary note.

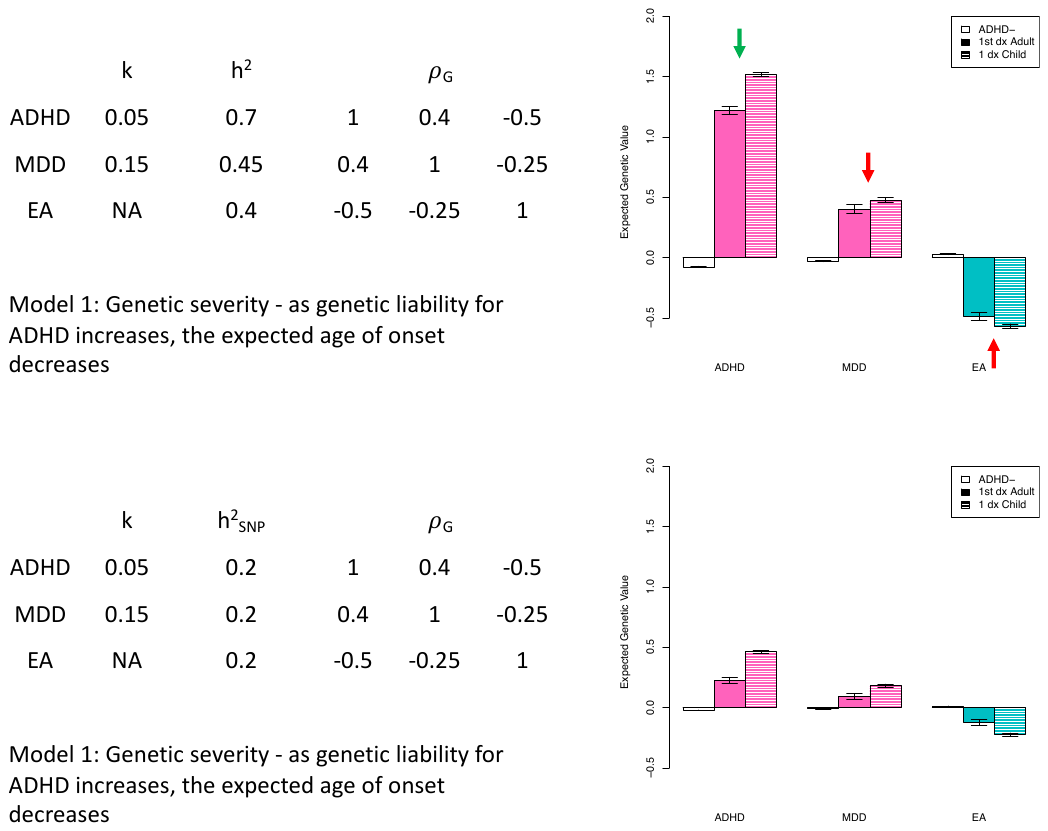

**Supplementary figure 12.** Adult diagnosis model 1: genetic severity. Here we use a multivariate, threshold liability model to generative illustrative genetic value profiles from a scenario that could underlie an adult versus child diagnosis of ADHD. These plots can be compared against main text Figure 4 to note qualitative differences (red arrows) or consistency (green arrows). On the left we show the population parameters that define the simulated population. Here, the age of diagnosis was chosen so a higher ADHD liability was associated with an earlier age of diagnosis. More methodological details and discussion are provided in the supplementary note.

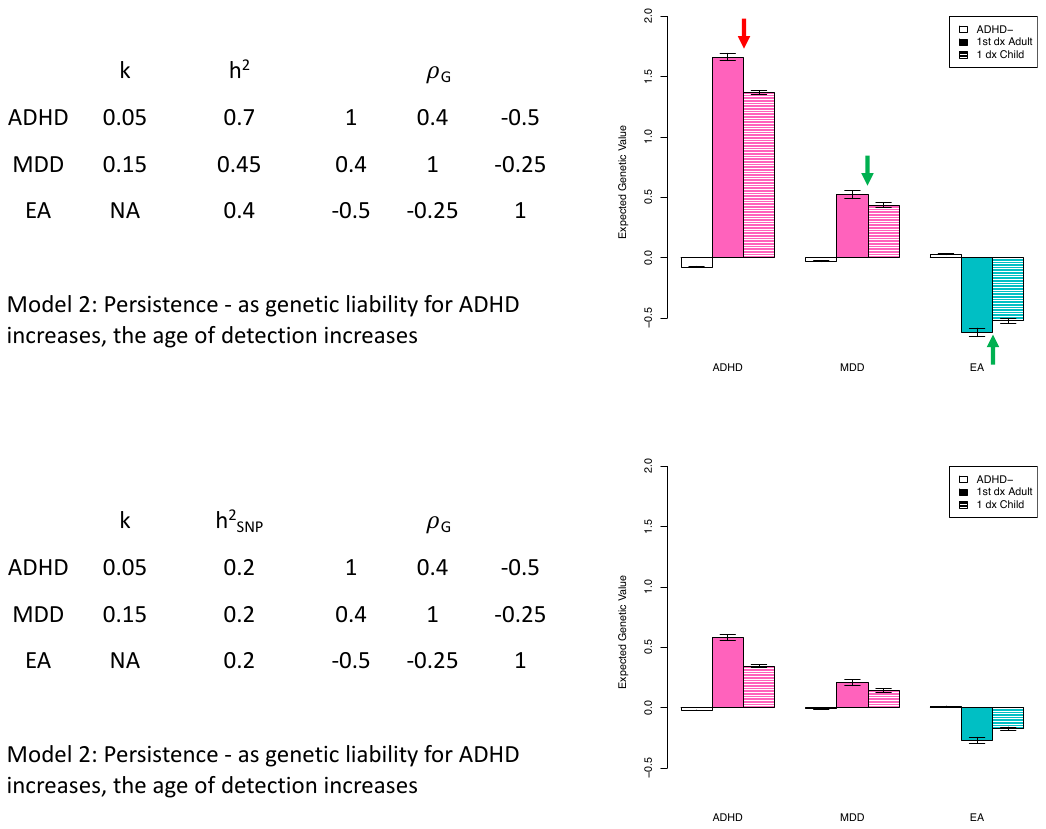

**Supplementary figure 13.** Adult diagnosis model 2: persistence. Here we use a multivariate, threshold liability model to generative illustrative genetic value profiles from a scenario that could underlie an adult versus child diagnosis of ADHD. These plots can be compared against main text Figure 4 to note qualitative differences (red arrows) or consistency (green arrows). On the left we show the population parameters that define the simulated population. Here, the age of diagnosis was chosen so a higher ADHD liability was associated with an later age of diagnosis. More methodological details and discussion are provided in the supplementary note.

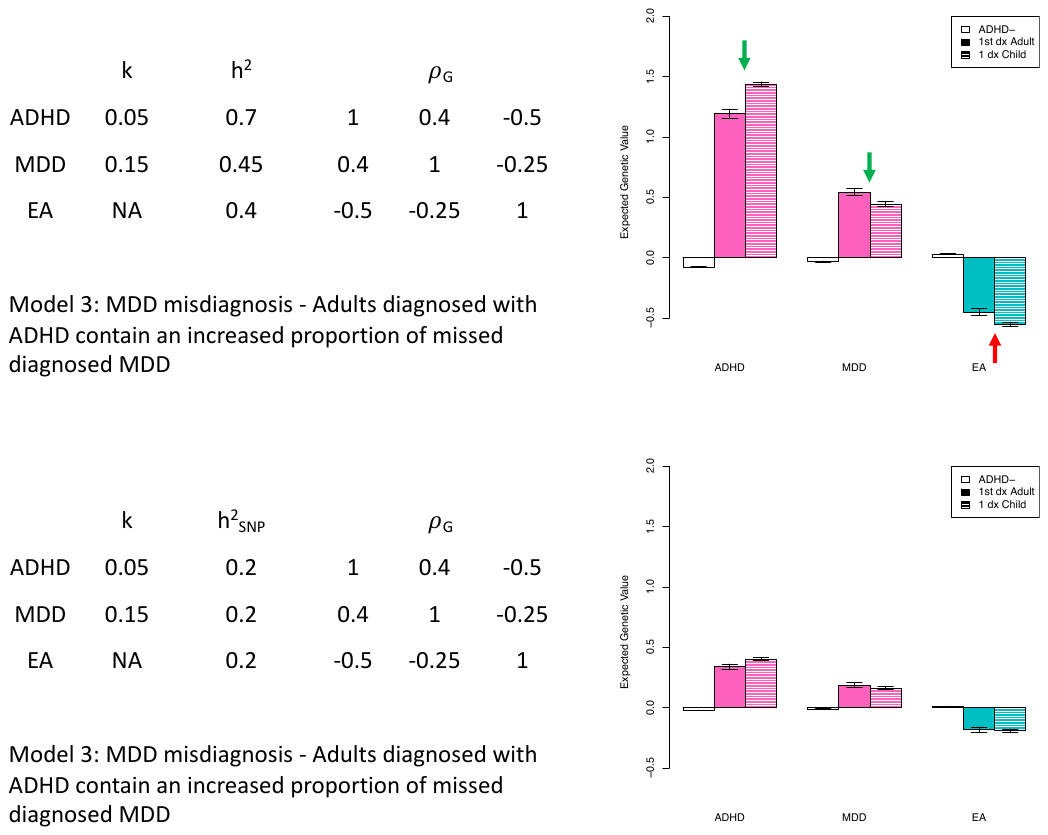

**Supplementary figure 14.** Adult diagnosis model 3: misdiagnosis. Here we use a multivariate, threshold liability model to generative illustrative genetic value profiles from a scenario that could underlie an adult versus child diagnosis of ADHD. These plots can be compared against main text Figure 4 to note qualitative differences (red arrows) or consistency (green arrows). On the left we show the population parameters that define the simulated population. Here, adult diagnosed ADHD was a mixture of ADHD (80%) and MDD (20%). More methodological details and discussion are provided in the supplementary note.

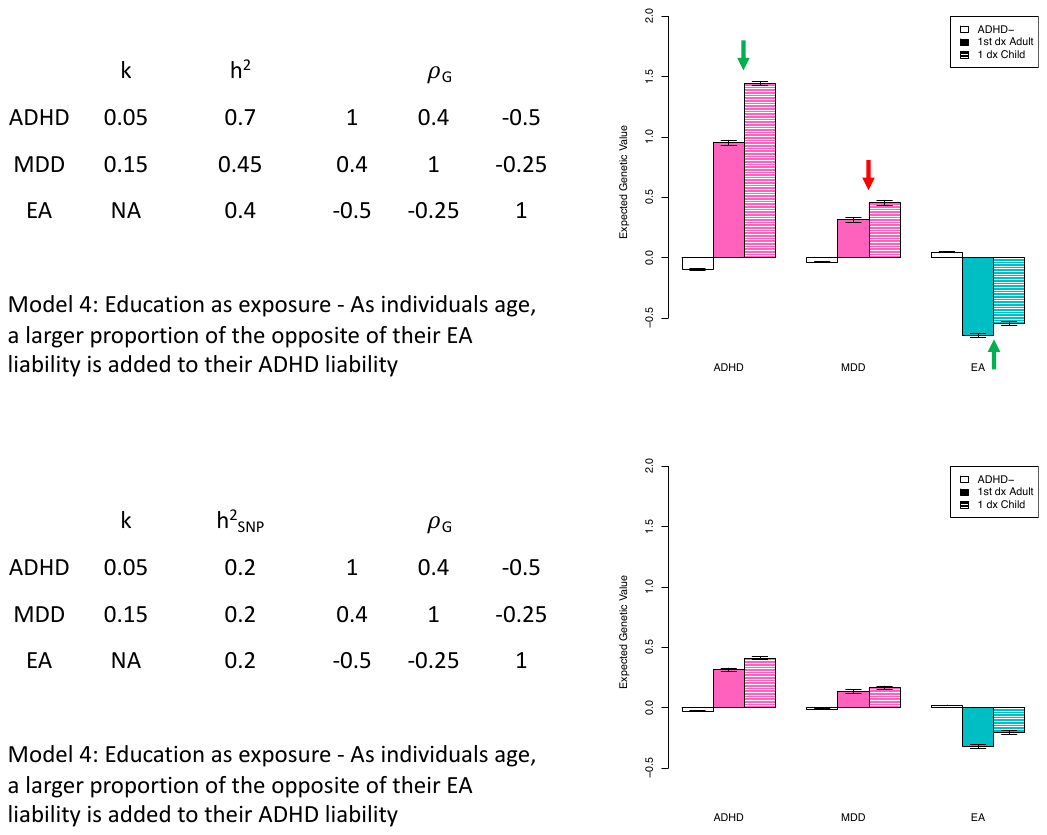

**Supplementary figure 15.** Adult diagnosis model 4: education as an exposure. Here we use a multivariate, threshold liability model to generative illustrative genetic value profiles from a scenario that could underlie an adult versus child diagnosis of ADHD. These plots can be compared against main text Figure 4 to note qualitative differences (red arrows) or consistency (green arrows). On the left we show the population parameters that define the simulated population. Here, adults diagnosed ADHD had the opposite of their EA liability score as a contribution to ADHD liability. More methodological details and discussion are provided in the supplementary note.

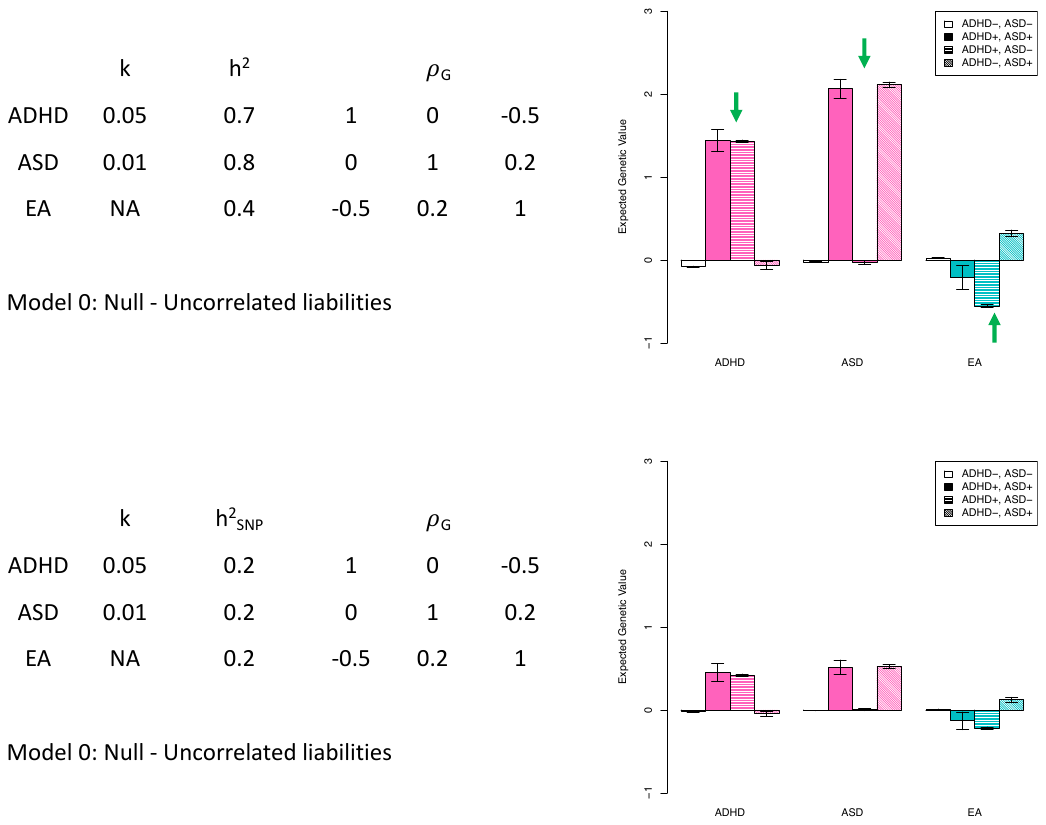

**Supplementary figure 16.** ADHD adjacent ASD model 0: uncorrelated liability. Here we use a multivariate, threshold liability model to generative illustrative genetic value profiles from a scenario that could underlie an ADHD adjacent ASD diagnosis. These plots can be compared against main text Figure 4 to note qualitative differences (red arrows) or consistency (green arrows). On the left we show the population parameters that define the simulated population. Here, ADHD and ASD arise from uncorrelated liability distributions, and those above the threshold for both receive both diagnoses. More methodological details and discussion are provided in the supplementary note.

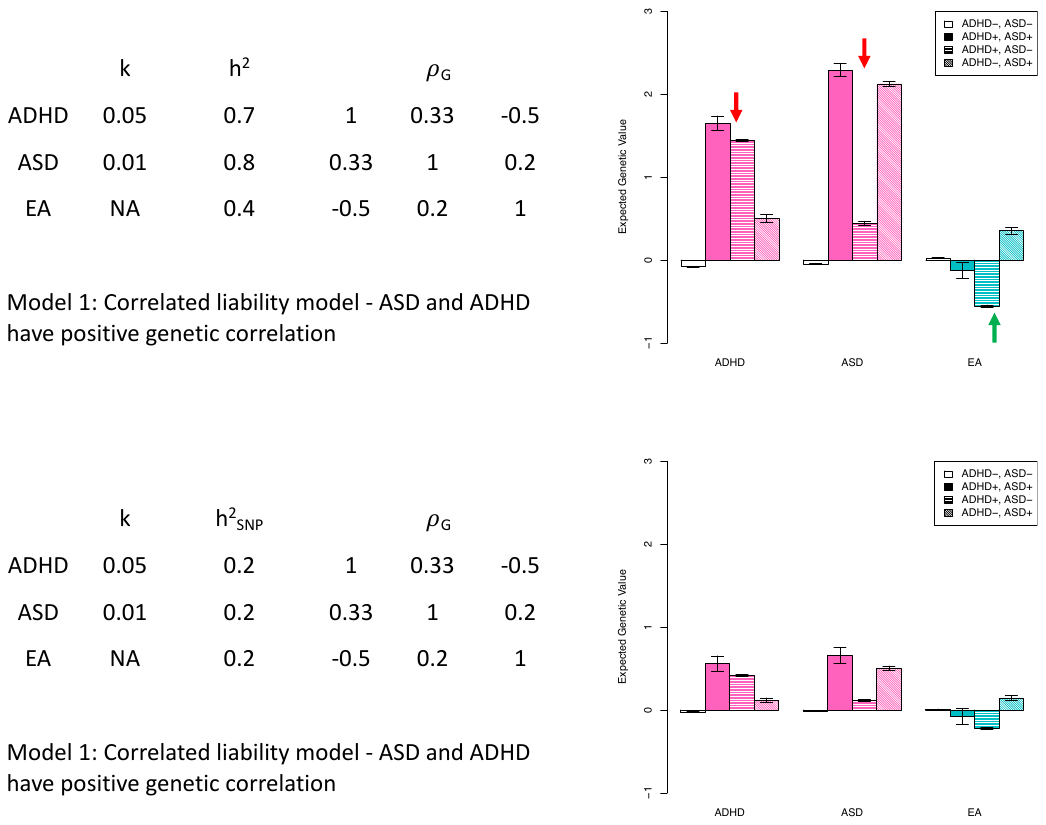

**Supplementary figure 17.** ADHD adjacent ASD model 1: correlated liability. Here we use a multivariate, threshold liability model to generative illustrative genetic value profiles from a scenario that could underlie an ADHD adjacent ASD diagnosis. These plots can be compared against main text Figure 4 to note qualitative differences (red arrows) or consistency (green arrows). On the left we show the population parameters that define the simulated population. Here, ADHD and ASD arise from correlated liability distributions, and those above the threshold for both receive both diagnoses. More methodological details and discussion are provided in the supplementary note.

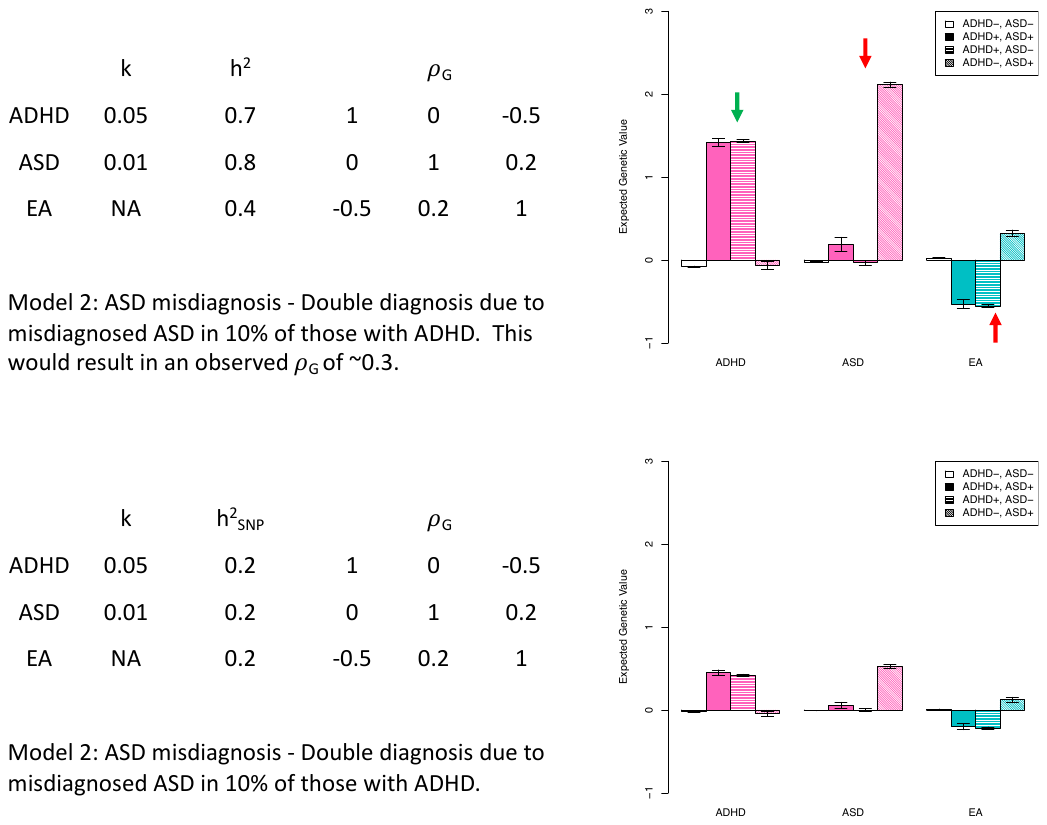

**Supplementary figure 18.** ADHD adjacent ASD model 2: ASD misdiagnosis. Here we use a multivariate, threshold liability model to generative illustrative genetic value profiles from a scenario that could underlie an ADHD adjacent ASD diagnosis. These plots can be compared against main text Figure 4 to note qualitative differences (red arrows) or consistency (green arrows). On the left we show the population parameters that define the simulated population. Here, in addition to those above the threshold for both disorders arising from uncorrelated liabilities, 10% of ADHD cases were assigned an adjacent ASD diagnosis. More methodological details and discussion are provided in the supplementary note.

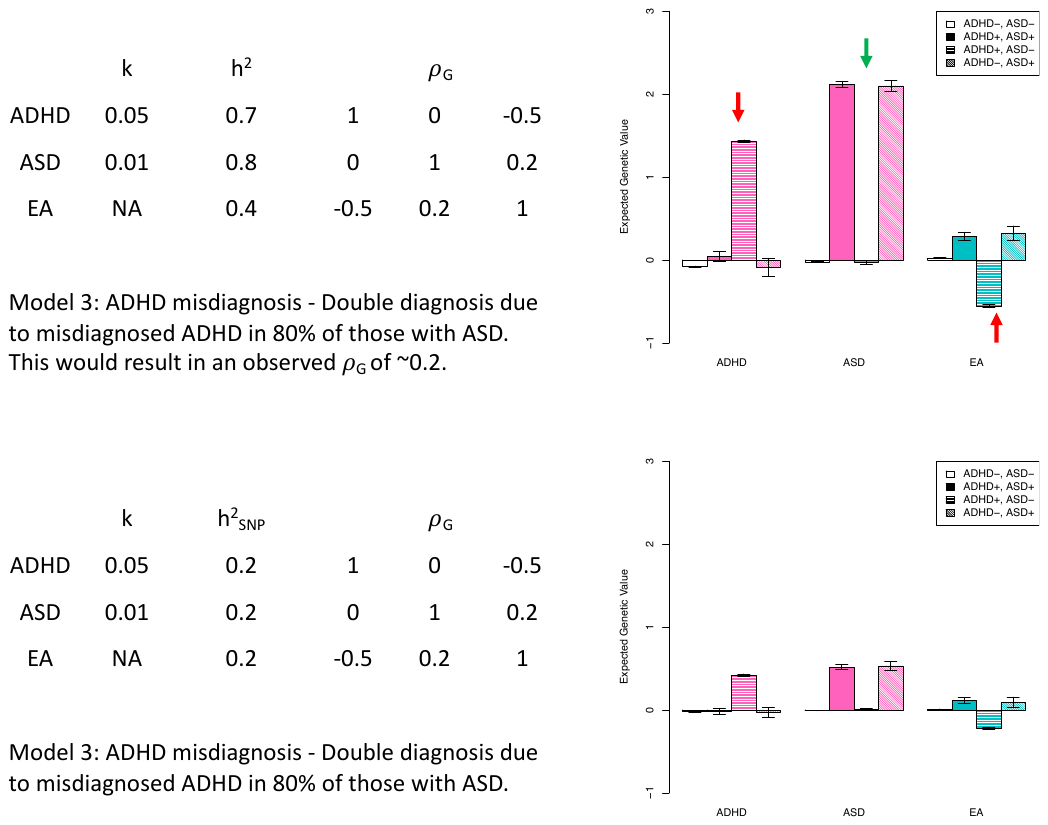

**Supplementary figure 19.** ADHD adjacent ASD model 3: ADHD misdiagnosis. Here we use a multivariate, threshold liability model to generative illustrative genetic value profiles from a scenario that could underlie an ADHD adjacent ASD diagnosis. These plots can be compared against main text Figure 4 to note qualitative differences (red arrows) or consistency (green arrows). On the left we show the population parameters that define the simulated population. Here, in addition to those above the threshold for both disorders arising from uncorrelated liabilities, 80% of ASD cases were assigned an adjacent ADHD diagnosis. More methodological details and discussion are provided in the supplementary note.

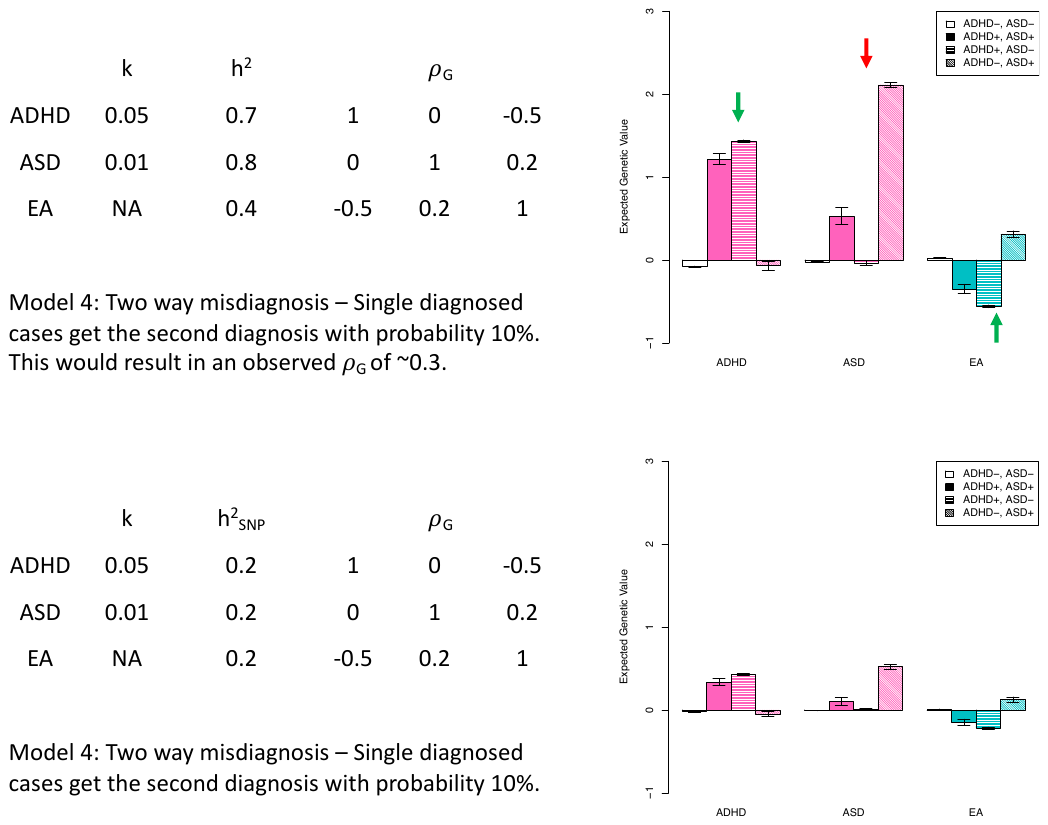

**Supplementary figure 20.** ADHD adjacent ASD model 4: two-way misdiagnosis. Here we use a multivariate, threshold liability model to generative illustrative genetic value profiles from a scenario that could underlie an ADHD adjacent ASD diagnosis. These plots can be compared against main text Figure 4 to note qualitative differences (red arrows) or consistency (green arrows). On the left we show the population parameters that define the simulated population. Here, in addition to those above the threshold for both disorders arising from uncorrelated liabilities, 10% of ASD cases were assigned an adjacent ADHD diagnosis and 10% of ADHD cases were assigned an adjacent ASD diagnosis. More methodological details and discussion are provided in the supplementary note.

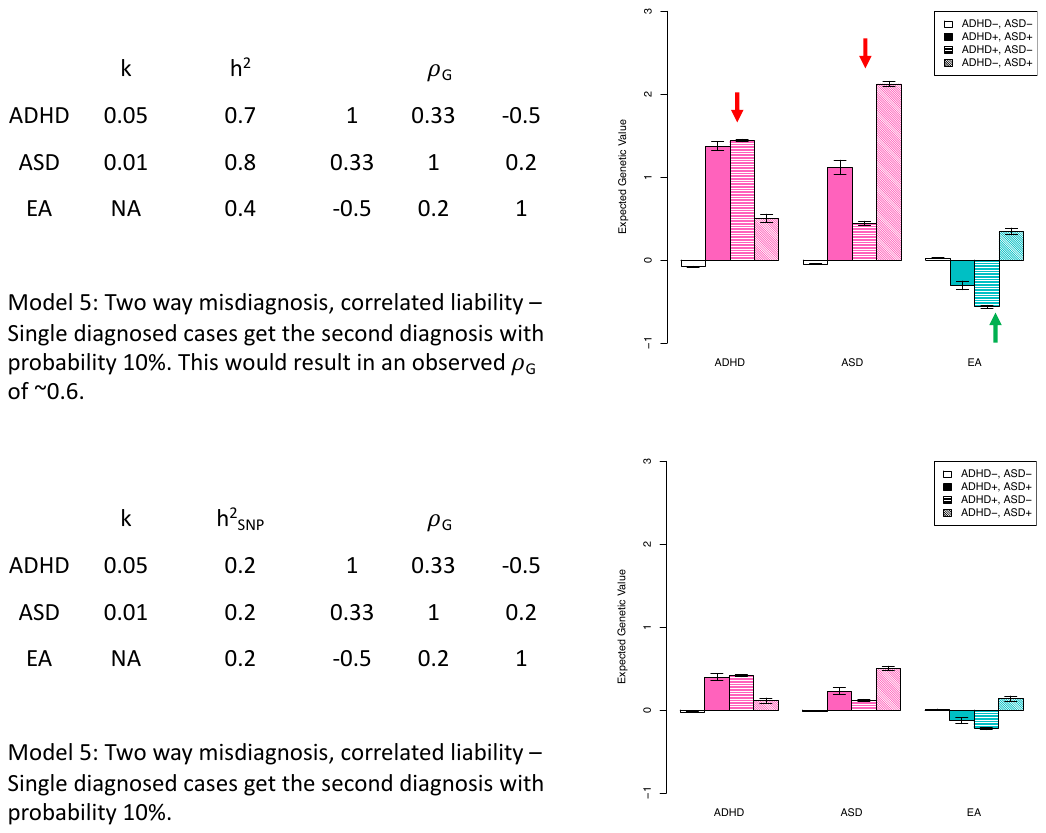

**Supplementary figure 21.** ADHD adjacent ASD model 5: two-way misdiagnosis, correlated liability. Here we use a multivariate, threshold liability model to generative illustrative genetic value profiles from a scenario that could underlie an ADHD adjacent ASD diagnosis. These plots can be compared against main text Figure 4 to note qualitative differences (red arrows) or consistency (green arrows). On the left we show the population parameters that define the simulated population. Here, in addition to those above the threshold for both disorders arising from correlated liabilities, 10% of ASD cases were assigned an adjacent ADHD diagnosis and 10% of ADHD cases were assigned an adjacent ASD diagnosis. More methodological details and discussion are provided in the supplementary note.

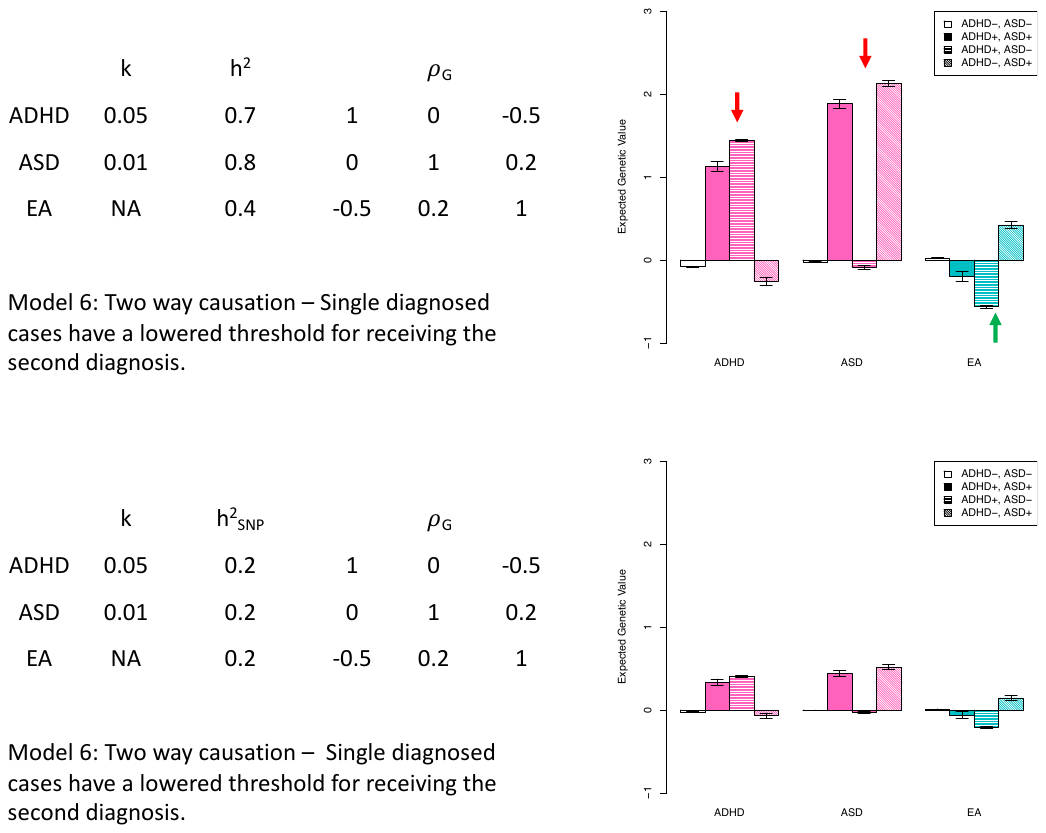

**Supplementary figure 22.** ADHD adjacent ASD model 6: two-way causation. Here we use a multivariate, threshold liability model to generative illustrative genetic value profiles from a scenario that could underlie an ADHD adjacent ASD diagnosis. These plots can be compared against main text Figure 4 to note qualitative differences (red arrows) or consistency (green arrows). On the left we show the population parameters that define the simulated population. Here, in addition to those above the threshold for both disorders arising from uncorrelated liabilities, ASD cases were potentially assigned an adjacent ADHD diagnosis on the basis of a lowered ADHD liability threshold and ADHD cases were potentially assigned an adjacent ASD diagnosis on the basis of a lowered ADHD liability threshold. More methodological details and discussion are provided in the supplementary note.

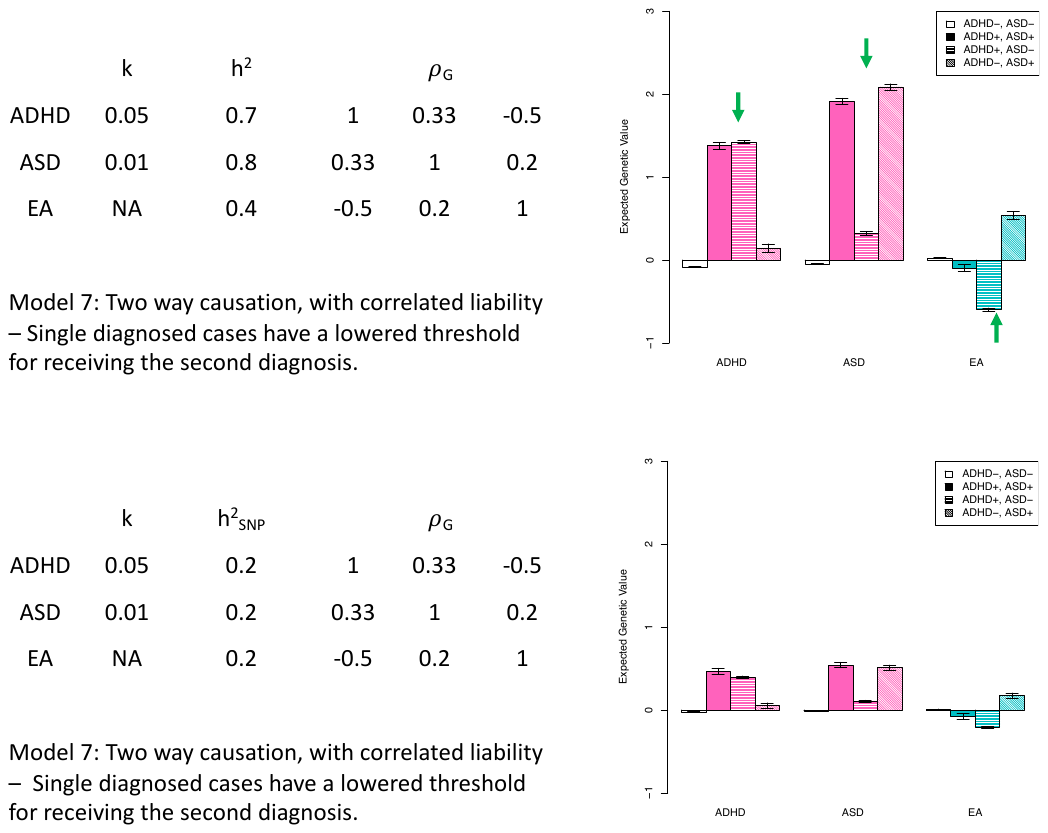

**Supplementary figure 23.** ADHD adjacent ASD model 7: two-way causation, correlated liability. Here we use a multivariate, threshold liability model to generative illustrative genetic value profiles from a scenario that could underlie an ADHD adjacent ASD diagnosis. These plots can be compared against main text Figure 4 to note qualitative differences (red arrows) or consistency (green arrows). On the left we show the population parameters that define the simulated population. Here, in addition to those above the threshold for both disorders arising from correlated liabilities, ASD cases were potentially assigned an adjacent ADHD diagnosis on the basis of a lowered ADHD liability threshold and ADHD cases were potentially assigned an adjacent ASD diagnosis on the basis of a lowered ADHD liability threshold. More methodological details and discussion are provided in the supplementary note.

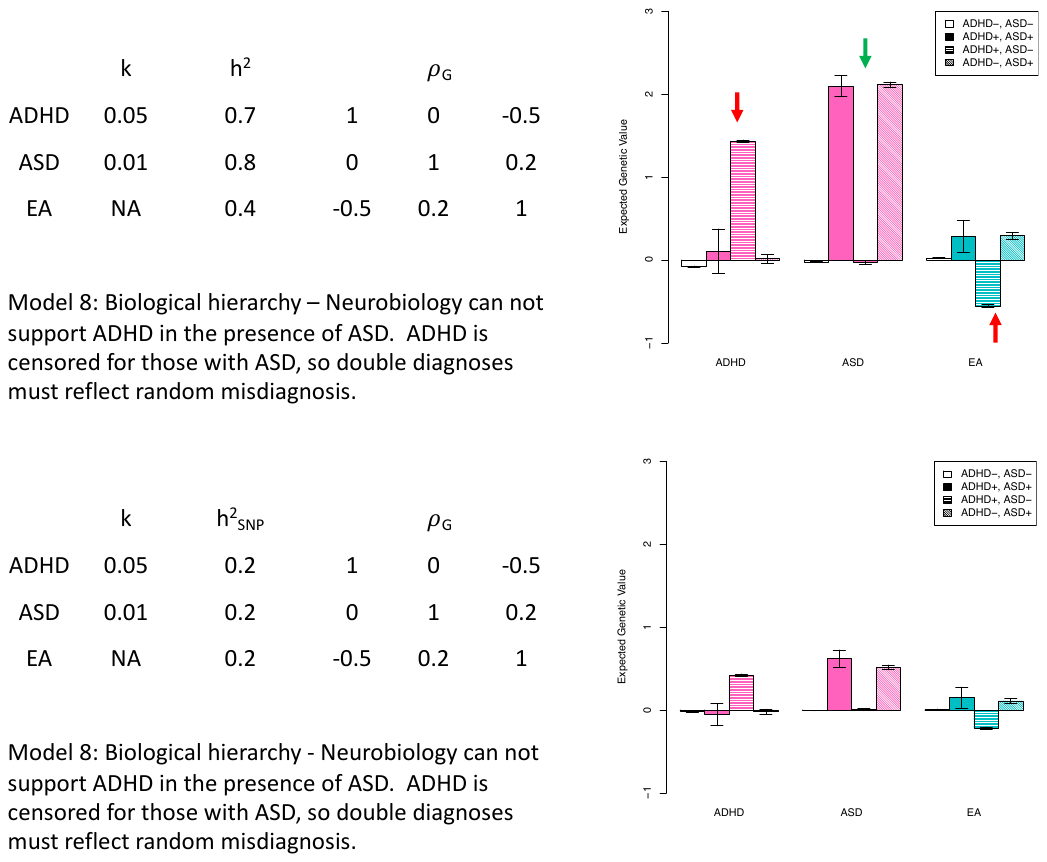

**Supplementary figure 24.** ADHD adjacent ASD model 8: biological hierarchy. Here we use a multivariate, threshold liability model to generative illustrative genetic value profiles from a scenario that could underlie an ADHD adjacent ASD diagnosis. These plots can be compared against main text Figure 4 to note qualitative differences (red arrows) or consistency (green arrows). On the left we show the population parameters that define the simulated population. Here, ASD cases above the ADHD threshold had their ADHD diagnosis removed. To create dual diagnosed individuals, a random 5% of ASD cases were assigned an ADHD diagnosis. More methodological details and discussion are provided in the supplementary note.

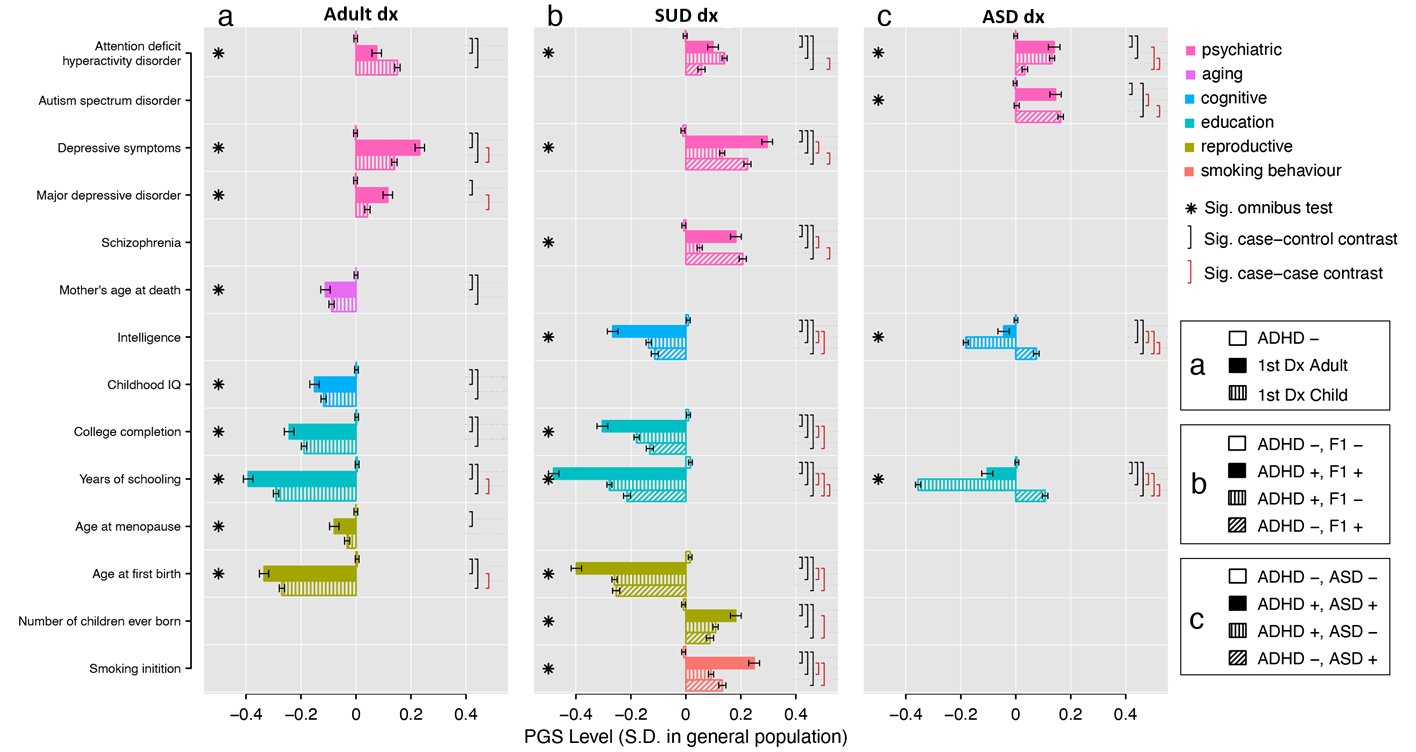

**Supplementary figure 25**.

Polygenic score (PGS) level in standard divisions (S.D.) to the general population (x-axis). PGS of fourteen brain and behavioral traits in six categories (y-axis). Except *Attention deficit hyperactivity disorder* and *Autism spectrum disorder,* the traits were significantly correlated with at least of one of the three ADHD-adjacent traits. **a.** Individuals not diagnosed with ADHD (ADHD -), first diagnosed with ADHD as an adult (1^st^ Dx Adult), and first diagnosed with ADHD as a child (1^st^ Dx Child), **b.** individuals diagnosed with neither ADHD, nor SUD (ADHD-, SUD-), both ADHD and SUD (ADHD+, SUD+), ADHD but not SUD (ADHD+, SUD-), and SUD but not ADHD (ADHD-, SUD+), and **c.** individuals diagnosed with neither ADHD, nor ASD (ADHD-, ASD-), both ADHD and ASD (ADHD+, ASD+), ADHD but not ASD (ADHD+, ASD-), and ASD but not ADHD (ADHD-, ASD+), Association of PGS to subgroups within each ADHD-adjacent trait were tested in single multinomial logistic regression. Significance level reported if p-value < 5x10^-8^. See Supplementary tables 14-16 for numeric values of single logistic regression.

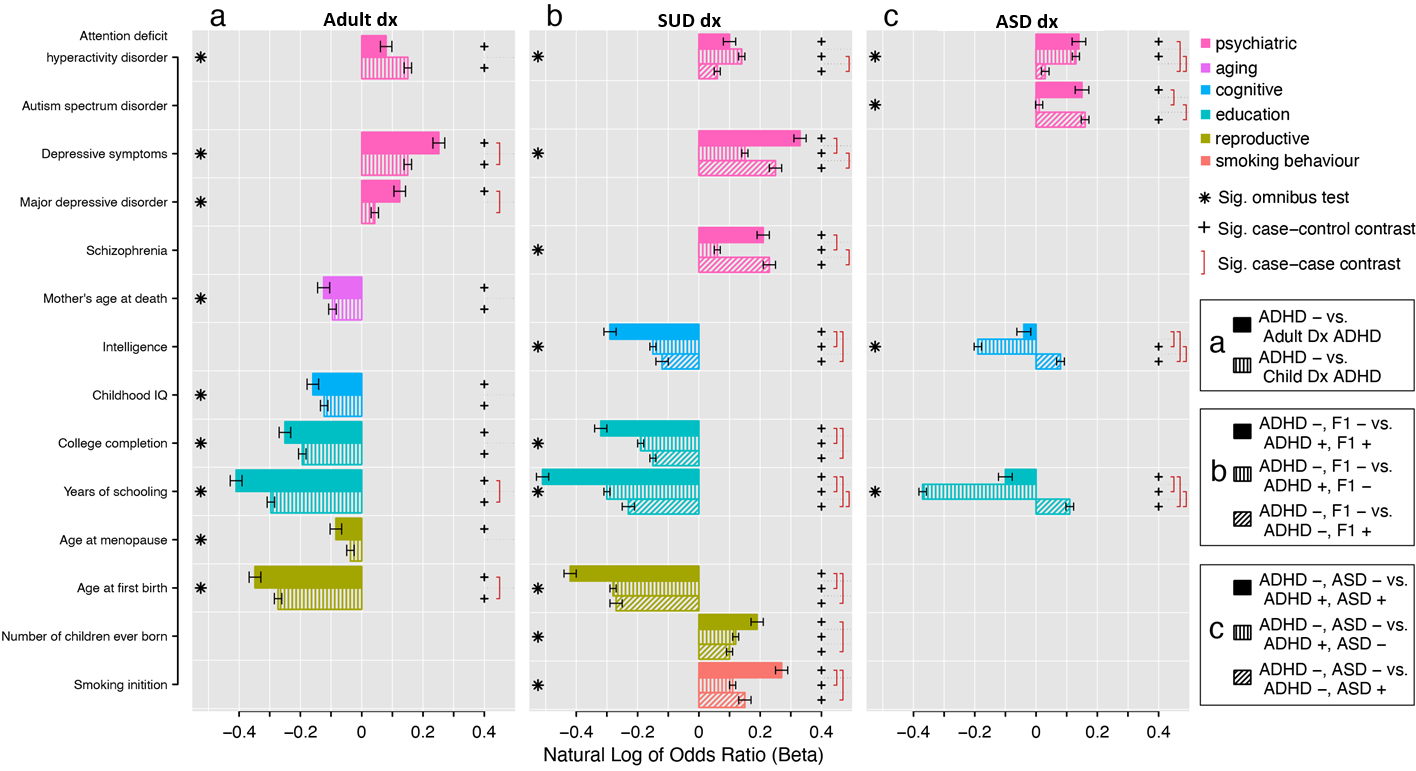

**Supplementary figure 26**.

PGS of fourteen brain and behavioral traits in six categories (y-axis). Except *Attention deficit hyperactivity disorder* and *Autism spectrum disorder,* the traits were significantly correlated with at least of one of the three ADHD-adjacent traits **a.** Individuals not diagnosed with ADHD (ADHD -), first diagnosed with ADHD as an adult (1^st^ Dx Adult), and first diagnosed with ADHD as a child (1^st^ Dx Child), **b.** individuals diagnosed with neither ADHD, nor SUD (ADHD-, SUD-), both ADHD and SUD (ADHD+, SUD+), ADHD but not SUD (ADHD+, SUD-), and SUD but not ADHD (ADHD-, SUD+), and **c.** individuals diagnosed with neither ADHD, nor ASD (ADHD-, ASD-), both ADHD and ASD (ADHD+, ASD+), ADHD but not ASD (ADHD+, ASD-), and ASD but not ADHD (ADHD-, ASD+), Association of PGS to subgroups within each ADHD-adjacent trait were tested in univariate multinomial logistic regression. Natural Log of Odds Ratio (beta) for subgroups of each ADHD-adjacent trait reported (x-axis) and significance level reported if p-value < 5x10^-8^. See Supplementary tables 14-16 for numeric values of single logistic regression.

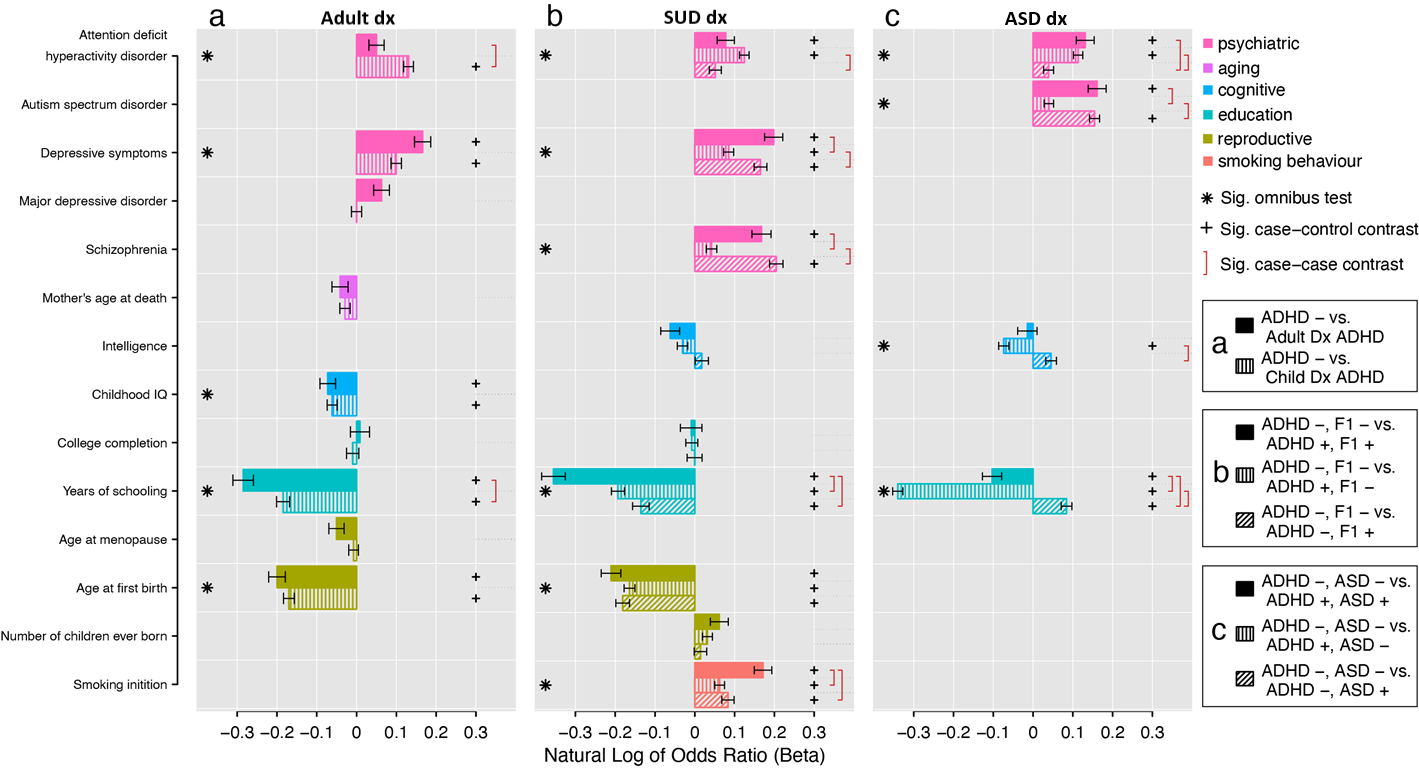

**Supplementary figure 27**.

PGS of fourteen brain and behavioral traits in six categories (y-axis). Except *Attention deficit hyperactivity disorder* and *Autism spectrum disorder,* the traits were significantly correlated with at least of one of the three ADHD variables. **a.** Individuals not diagnosed with ADHD (ADHD -), first diagnosed with ADHD as an adult (1^st^ Dx Adult), and first diagnosed with ADHD as a child (1^st^ Dx Child), **b.** individuals diagnosed with neither ADHD, nor SUD (ADHD-, SUD-), both ADHD and SUD (ADHD+, SUD+), ADHD but not SUD (ADHD+, SUD-), and SUD but not ADHD (ADHD-, SUD+), and **c.** individuals diagnosed with neither ADHD, nor ASD (ADHD-, ASD-), both ADHD and ASD (ADHD+, ASD+), ADHD but not ASD (ADHD+, ASD-), and ASD but not ADHD (ADHD-, ASD+), Association of PGS to subgroups within each ADHD-adjacent trait were tested in joint multinomial logistic regression. Natural Log of Odds Ratio(beta) for subgroups of each ADHD-adjacent traits reported (x-axis) and significance level reported if p-value < 5x10^-8^. See supplementary tables 11-13 for numeric values of joint multinomial logistic regression.
